# Organ damage proteomic signature identifies patients with MASLD at-risk of systemic complications

**DOI:** 10.1101/2024.10.28.24316149

**Authors:** Carlos José Pirola, Luis Diambra, Tomas Fernández Gianotti, Gustavo Osvaldo Castaño, Julio San Martino, Martin Garaycoechea, Silvia Sookoian

## Abstract

**Background & objectives:** Metabolic steatotic liver disease (MASLD), which affects more than 30% of the world’s population, is associated with multisystemic comorbidities. We combined multidimensional OMICs approaches to explore the feasibility of using high-throughput targeted circulating proteomics to track systemic organ damage and to infer its underlying mechanisms at the molecular level.

**Desing:** We tested a 92-plex panel of prioritised proteins with pathophysiological relevance to organ damage in serum samples of patients with in-depth phenotyping. We included proteomics data from 60,042 people in the discovery and validation stages. The identified protein was validated across diverse study designs and cross-proteomic platforms. Deconvolution strategies were used to investigate whether the affected liver is involved in expressing biomarkers of organ damage. To assess the cell type-specific transcriptional changes of the selected target we used liver organoid data.

**Results:** The implicated proteins, including ADGRG1 (GPR56), are deregulated in patients with MASLD at-risk of progressive disease and significant fibrosis. ADGRG1 liver expression profile mirrors the circulation pattern. ADGRG1 levels were associated with increased risk of end-stage liver disease and a modest but clinically significant risk of death, chronic obstructive pulmonary disease, and ischaemic heart disease over 16 years of follow-up. Mechanistic insight shows that ADGRG1 shifts its transcriptional profile from low expression to upregulation in liver cells of the fibrotic niche in response to injury.

**Conclusions:** Our study provides a framework of potential mechanisms associated with systemic organ injury that facilitates holistic management by stratifying patients with MASLD into subclasses at-risk of extrahepatic manifestations.

## INTRODUCTION

More than 30% of the world’s population is affected by metabolic dysfunction-associated steatotic liver disease (MASLD) [1] [2]. This complex disorder arises from interactions with various cardiometabolic [3] [4], environmental [5] and genetic risk factors [6]. The histological expression of MASLD has several phenotypic stages. These include metabolic dysfunction-associated steatosis (MASL) and metabolic dysfunction-associated steatohepatitis (MASH), the latter with varying degrees of fibrosis that can lead to cirrhosis and hepatocellular carcinoma (HCC) [7] [4]. The natural history and histological evolution of MASLD stages are dynamic [8], causing interindividual phenotypic variability. However, certain histological features, including hepatocellular ballooning, portal inflammation, and fibrosis stage play a crucial role in determining the disease trajectory [8]. Numerous epidemiological studies have demonstrated that the disease’s natural history is significantly modified by the presence of significant or advanced fibrosis [9] [10] [11] [12] [13].

Besides, MASLD has been associated with damage or dysfunction in other organs of the body, leading to conditions such as cardiovascular disease (CVD) [14] [15] [16] [17] [18] [19], chronic kidney disease [20] [21], obstructive sleep apnoea [22], and non-liver cancers [23] among many other systemic manifestations [24]. MASLD is also associated with increased all-cause mortality in a dose-dependent manner across the disease stages, which appears to be mainly due to cirrhosis, HCC and extrahepatic cancers, and CVD [25].

The liver is responsible for producing and secreting most of the abundant plasma proteins [26]. Significant progress has been made in identifying proteomic signatures associated with patients with MASH at-risk of the most severe liver outcomes [27–32]. However, the disease mechanisms behind the systemic organ damage associated with MASLD remain poorly understood. Furthermore, it is unclear whether the liver is responsible for producing proteins involved in systemic complications and whether patients with MASLD and advanced disease are more likely to develop severe disease-related multisystemic complications.

Here, we hypothesized that progressive MASLD, characterized by inflammation and fibrosis but not steatosis alone, may predispose individuals to systemic organ damage and extrahepatic complications. Therefore, we combined multidimensional OMICs approaches to explore the feasibility of using targeted high-throughput circulating proteomics to track MASLD-associated systemic organ damage. In addition, we aimed to uncover mechanistic biomarkers that may serve to characterise the molecular subtypes of the disease that are associated with systemic complications.

## METHODS

### Patients, data collection and study design

Our hypothesis-driven study consisted of an integrative analysis of several OMICs approaches with clinical data, as shown in **Fig. 1**. The discovery phase involved the identification of proteins from the circulation using a 92-plex panel of prioritized signatures with pathophysiological importance in organ damage (Olink Proteomics, Uppsala, Sweden); see **Table S1** for proteins list and **Fig.S1** for disease area and associated biological process. This phase included 88 serum samples from individuals with MASLD diagnosed by in-depth clinical and histological phenotyping, matched for age and sex, and representing the full histological disease spectrum and control subjects (**Table 1**; **Table S2**). Patients included in the discovery cohort are provided with prospective information on the occurrence of extrahepatic comorbidities, as well as mortality data during the subsequent follow-up period (lasting between two and eight years from the date of the baseline biopsy and samples collection).

**Figure 1.**
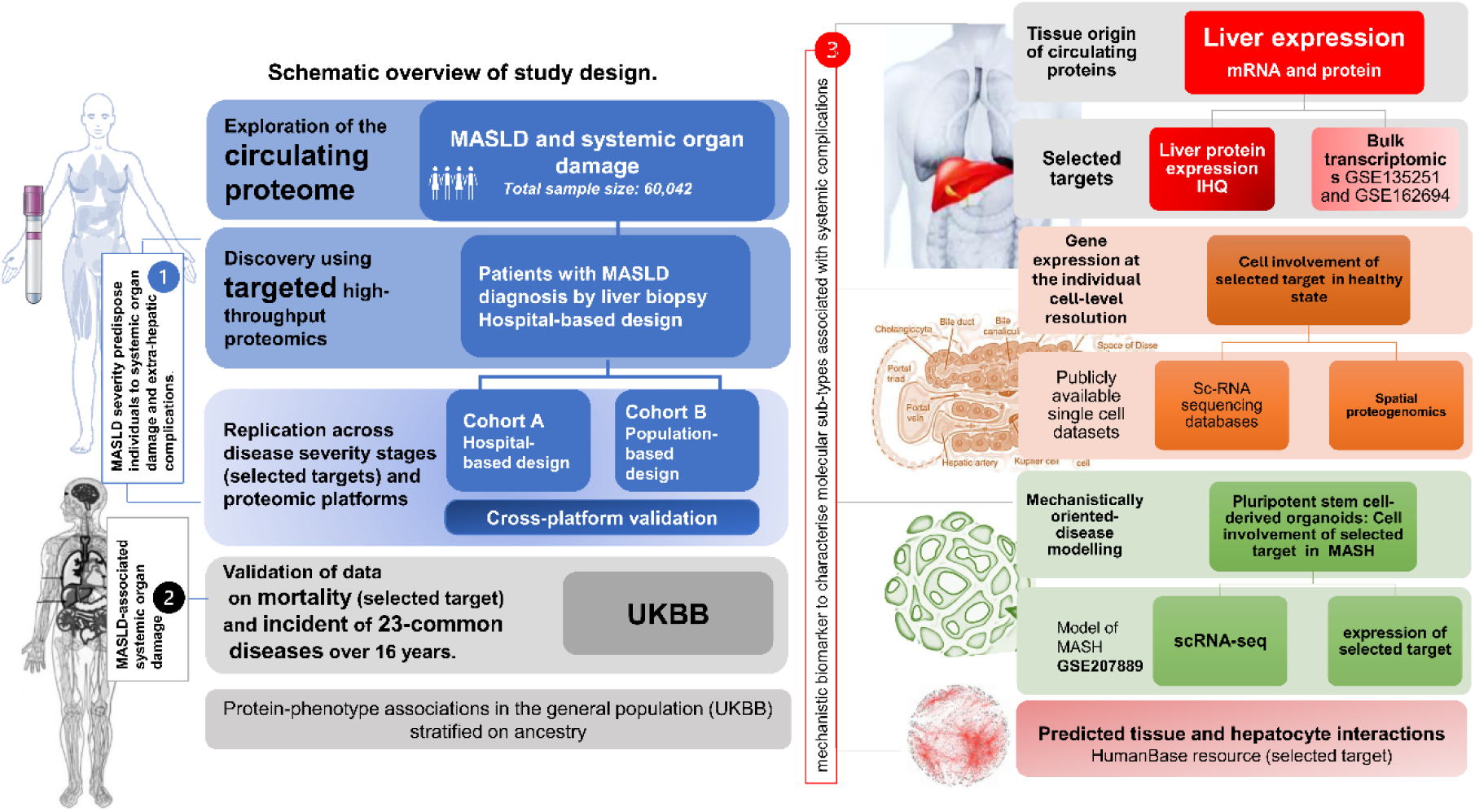
Workflow and study phases. Study design and workflow of sample processing, data generation, and integrative analysis of phases data. 1-3: Research questions and hypothesis.

**Table 1:**
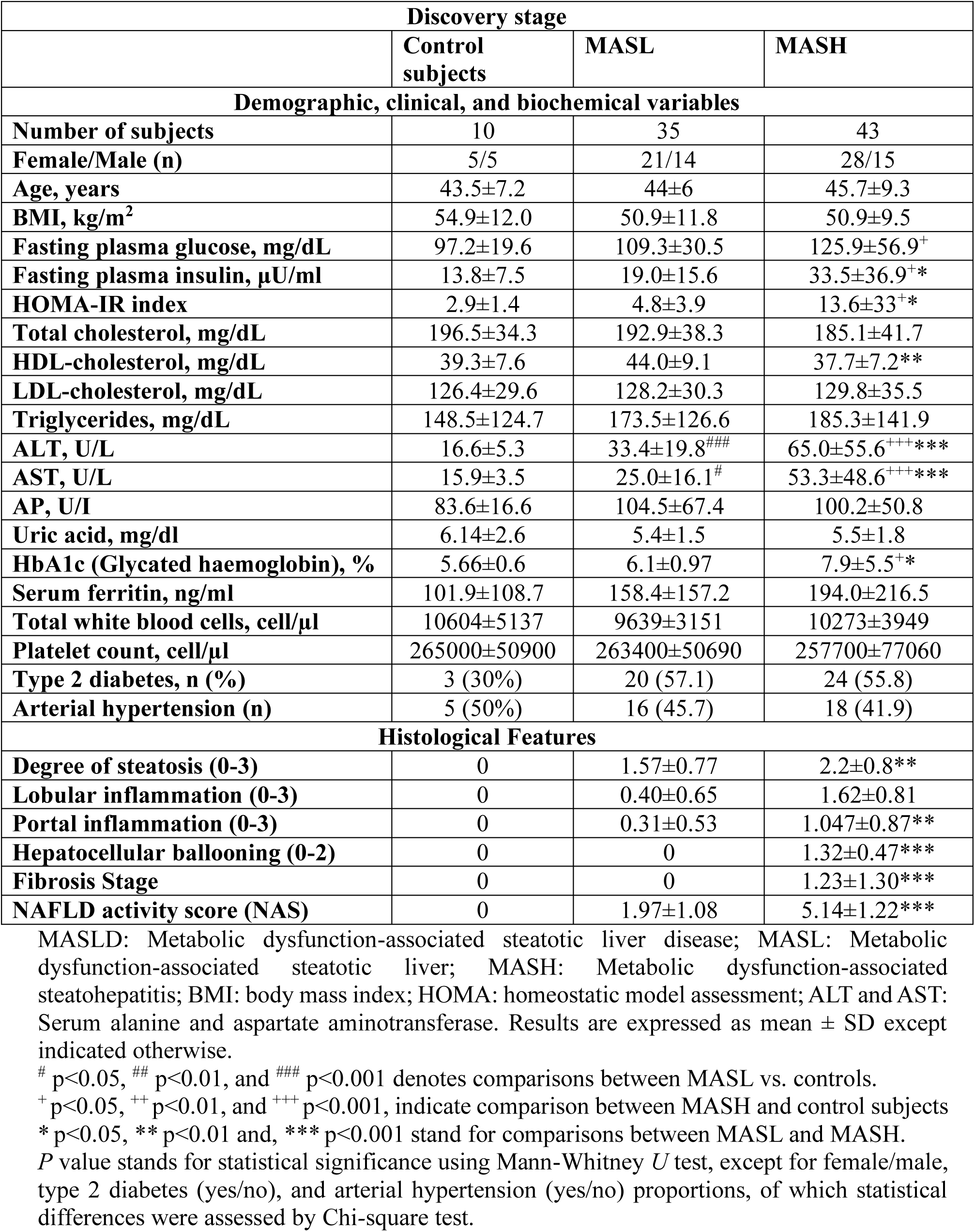
Baseline patients characteristics in the discovery cohort.

The replication phase comprised a focused examination of proteomic data identified from the discovery stage, encompassing two cohorts of individuals of European ancestry with distinct study designs and utilising publicly available data for fair reuse. This stage was designed to provide insight into whether the identified proteins in the first stage are more prevalent in the circulation of patients with more severe disease. The replication cohort A comprised retrievable metadata of 191 plasma samples [28] of patients from the hospital-based European NAFLD Registry [33], processed for proteomics using the SomaScan v.4.0 platform (Soma-Logic, Inc.) and diagnosed by liver biopsy. The replication cohort B comprised proteomics data retrieved from 38,890 population-based European-ancestry participants from the UK Biobank (UKBB) and processed for proteomics using the Olink Explore-1536 platform [32]. In addition, the replication cohort B included proteomics data retrieved from 20,873 Icelanders that were measured with the SomaScan v.4 assay [32]. A total of 610 cases with MASLD and 262 cases with cirrhosis were identified in the UKBB population, along with 181 cases with a diagnosis of MASLD and 73 cases with cirrhosis in the Icelandic cohort. The replication cohorts were also used to investigate the consistency of selected target effects between the two different high-throughput proteomics platforms.

The phase of exploring and validating the longitudinal relationship between the selected biomarker and the incident of 23 common systemic diseases that are associated with disability, morbidity, and reduction of life expectancy and mortality, included retrievable data from ref. [34] and collected from the UKBB (*n*=47,600) over 16 years using the Olink proteomic platform. We also explored protein-phenotype associations in the general population of the UKBB by reusing proteomics data from published datasets [35].

Liver protein expression of the identified targets was assessed by immunohistochemistry in 31 patient biospecimens paired with plasma proteomic profiled samples to explore the tissue origin of circulating proteins (12 cases of formalin-fixed paraffin-embedded liver tissue for each biomarker). Additionally, we included transcriptomic analyses by examining liver bulk RNA-sequencing expression data from two external and independent sources in the NCBI Gene Expression Omnibus (GEO) repository that profiled gene expression in patients with MASLD at different stages of the disease severity (GSE135251 and GSE162694).

The phase of cell and spatially resolved expression of a selected protein in the healthy liver was assessed by exploring the spatial proteogenomic atlas available at https://www.livercellatlas.org/umap-humanAll.php.

To investigate the involvement of the selected target in cell type expression, we included a mechanistically oriented phase using pluripotent stem cells (hPSCs)-derived human liver organoids (HLOs) that recapitulate the transcriptional landscape of major liver cell types in a model that mimics the inflammatory and fibrotic injury in human MASH [36]. Additional details can be found in **Supplementary file**.

### Ethical approval

Biological specimens, including blood samples and liver biopsies, were collected from all subjects with written, informed consent under Institutional Review Board (Comité de Ética en Investigación (CEI) intervenient Hospital A Zubizarreta Buenos Aires, Argentina) approved protocols with protocol numbers: 104/HGAZ/09, 89/100, 1204/2012, and updated and aproved DI-2019-376-GCABA-HGAZ, DI-2019-376-GCABA-HGAZ. Protocol entitled “Biomarkers of systemic organ damage” was registered with GCBA (Government of the City of Buenos Aires Argentina) protection data PRIISA BA (“Plataforma de Registro Informatizado de Investigaciones en Salud de Buenos Aires”-“Buenos Aires Health Research Computerized Registry Platform”): ethical approval was given under the reference number 8987. All data were de-identified prior to use in the study. All investigations were performed in accordance with the guidelines set forth in the 1975 Declaration of Helsinki, as revised in 1993.

### Patient and Public Involvement

It was not appropriate or possible to involve patients or the public in the design, or conduct, or reporting, or dissemination plans of our research

### Statistics

Continuous variables are expressed as means ± SD, and categorical variables were expressed as numbers and percentages. The Mann-Whitney U test was used to compare clinical, biochemical and histological characteristics, except for the female/male ratio, frequency of type 2 diabetes and arterial hypertension between the groups studied, which were evaluated using the chi-squared test. For more than two group comparison, statistical analysis was conducted using the nonparametric Kruskal-Wallis test followed by Dunn test. In the case of adjusted for covariates analyses, we used either ANOVA, linear or logistic regression after logarithmisation of variables non-normally distributed as assessed by Q-Q plots. A *p*-value less than 0.05 was considered statistically significant. Receiver operating characteristic (ROC) analyses and AUC calculations were performed by rocreg or roccomp subroutines implemented in STATA v16 (StataCorp LLC, College Station, TX, USA). When applicable, *p* values were adjusted using Benjamini-Hochberg to control for the false discovery rate (FDR) except indicated otherwise.

## RESULTS

### Biomarkers of organ damage in the discovery phase

The analysis of the circulating expression profile between samples of patients with MASL and MASH, processed for proteomics using the validated organ damage panel, revealed that four proteins exhibited differential upregulation in MASH after adjusting for relevant covariates (**Fig.2a-d**), including body mass index (BMI), age, sex, type 2 diabetes (T2D), and arterial hypertension. Logistic regression analysis showed that these proteins increased the risk of having MASH (ADGRG1, adhesion G-protein-coupled receptor G1, also known as GPR56: OR 2.63 95% CI 1.27-5.60 *p* =9.7e-03; MVK, mevalonate kinase: OR 1.78 95% CI 1.13-2.82, *p* =1.4e-02; NPPC, natriuretic peptide C: OR 1.74 95% CI 1.12-2.71, *p* =1.3e-02; and AGR2, anterior gradient 2, protein disulphide isomerase family member: OR 3.80 95% CI 1.38-10.50 *p* =1.0e-02).

**Figure 2:**
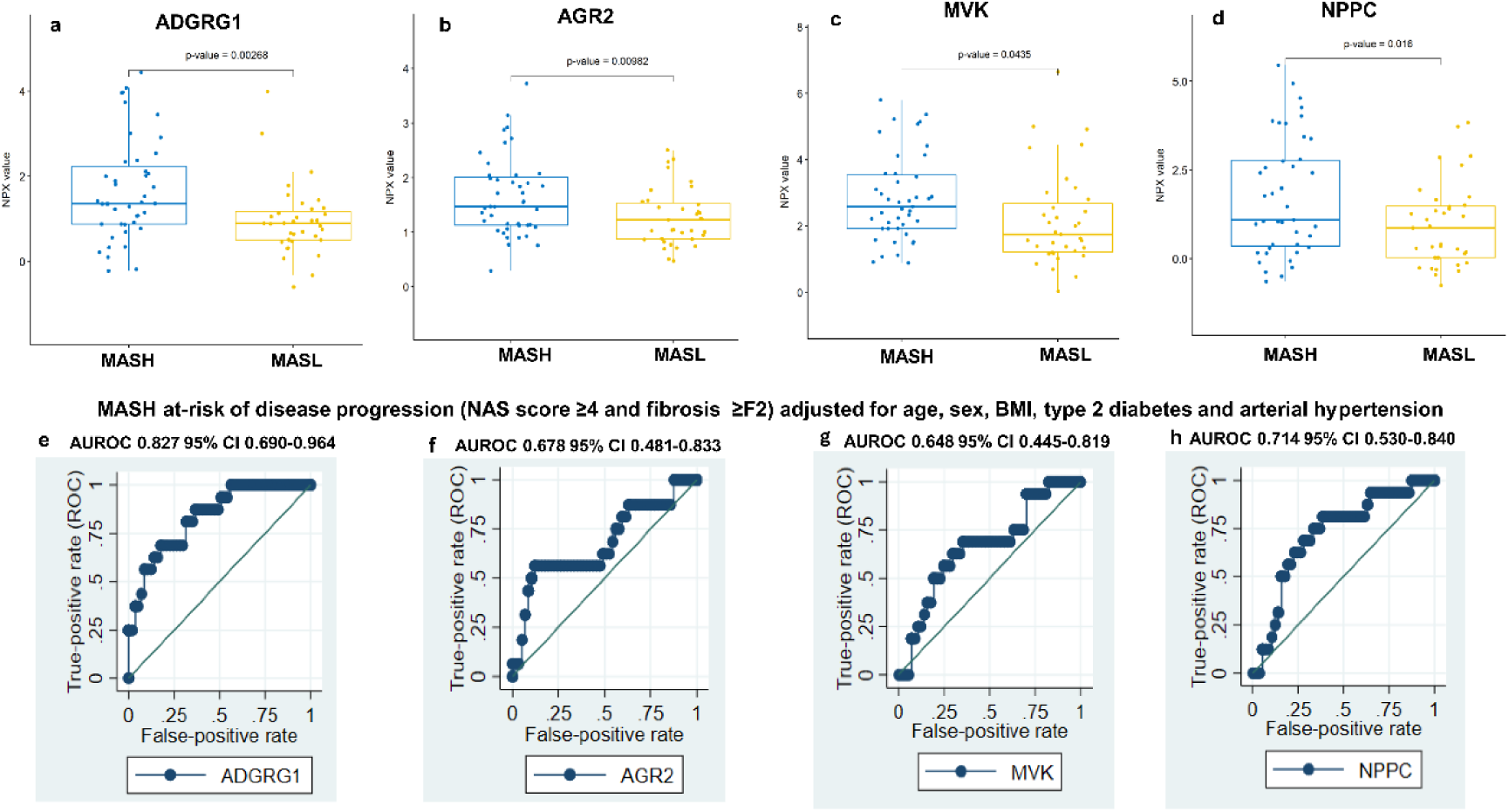
Selected targets deregulated in MASH at-risk of disease progression. **a-d**. A comparison of the circulating profile between MASL and MASH patient groups for four significantly deregulated proteins (ADGRG1, AGR2, MVK, and NPPC); *P* values are for t-test. **f-i**. AUROC performance of the selected deregulated targets in patients with MASH at-risk of disease progression according to NAS score ≥4 and fibrosis ≥F2. AUROCs are adjusted for age, BMI, type 2 diabetes and arterial hypertension.

When looking at the profile of patients with MASLD at-risk of disease progression, defined as having a NAFLD activity score–clinical research network (NAS) score ≥4 and fibrosis stage equal or higher than 2 [28,37], we found that ADGRG1 was the most significant protein associated with progressive MASH, *p=* <0.00001. ADGRG1 demonstrates an area under the receiver operating characteristic (AUROC) of 0.871 for at-risk MASH classification and an AUROC of 0.822 for MASH with significant fibrosis defined as fibrosis scores ≥2. The results adjusted for potential confounding variables, including age, sex, BMI, T2D, and arterial hypertension, remain statistically significant (at-risk AUROC 0.827 95% CI 0.690-0.964) (**Fig.2e-h**).

We also looked at the circulating pattern of the four above mentioned proteins according to the histological features associated with more severe disease. When stratifying patients based on portal inflammation, ranging from 0 to 3 and comparing 0 (none)-1 (mild) versus 2 (moderate)-3 (marked) we found ADGRG1 and NPPC to be differentially expressed in patients with higher scores (**Fig.S2a**). The comparison of patients with different degrees of lobular inflammation scored from 0-3 based upon the number of foci that show lobular inflammation *per* 20x fields showed that MVK and NPPC remain significantly deregulated (**Fig.2b**). A stratification of patients based on their fibrosis stage, ranging from 0 to 4, revealed that ADGRG1 remained significantly deregulated in the group of patients with liver fibrosis (**Fig.S2c**).

The differential abundance of these proteins in the circulation of subjects without MASLD (control group) was compared with that in subjects with MASL and MASH. ADGRG1 and MVK levels were found to be significantly increased in patients with MASH compared to the control group, after adjustment for the main covariates; detailed comparisons are shown in **Fig.S3**.

The analysis of outcomes in terms of mortality and extrahepatic comorbidities was based on records for patients with MASLD and was assessed according to the disease status, either MASL or MASH. Patients with MASH presented higher proportion of major outcomes (30.3%) compared to patients with MASL (2.94%), Chi-square *p* =0.01. The mortality rate for non-liver-related causes of death in patients with MASH was 8.82%, while in patients with MASL, it was 0%.

### Selected targets in the replication phase

The four proteins potentially involved in MASLD-related systemic organ injury were included in the replication phase, comprising ADGRG1, AGR2, MVK, and NPPC. In replication cohort A, results for differentially expressed soma probes after correction for sex, cohort centre and T2D were based on retrievable metadata of patients stratified as advanced-MASLD “no” (fibrosis F0-F2) versus advanced-MASLD “yes” (fibrosis score F3-F4) from ref. [28]. This dataset includes only information on ADGRG1 (GPR56), AGR2, and MVK. ADGRG1 was identified as the most robust circulating marker of organ damage, with significantly increased expression in patients with advanced disease, regardless of gender (**Fig.3a,b,c,j**). Plasma levels of AGR2 were significantly increased in patients with mild fibrosis. However, the results remain significant only in men (**Fig.3d,e,f,j**). MVK protein levels did not differ significantly between groups (**Fig.3g,h,i,j**). The adjusted protein levels for sex and for the presence of T2D indicates that ADGRG1 classifies patients with advanced MASLD with an AUROC of 0.677 (**Fig3j**).

In replication cohort B, results for differentially expressed proteins showed that the four proteins were associated with MASLD compared to the control (healthy) population. However, ADGRG1 and MVK were the most significantly associated biomarkers of organ damage in subjects with MASLD compared to the UKBB control population (**Fig.3m**). Likewise, ADGRG1 showed the most significant effects in MASLD compared to the non-MASLD population among Icelanders (**Fig.3n**). In the analysis of protein levels in patients diagnosed with cirrhosis compared to the general population, the only protein that remains significantly associated with more robust effects in both cohorts and measured by both proteomic platforms, OLINK and SomaScan v.4.0 was ADGRG1 (**Fig.3o,p)**. Levels of NPPC and MVK were significantly increased in patients with cirrhosis in the UKBB dataset but not among Icelanders (**Fig.3o,p**).

When comparing protein levels in patients with cirrhosis vs MASLD without cirrhosis, there were clear differences, and proteins that were significantly associated with MASLD not only lost significance when compared to cirrhosis, but also may show an inverse effect, as in the case of MVK (**Fig.3q,r**). However, for plasma ADGRG1 levels, there was a high degree of concordance in the direction of effects between the two proteomic datasets with a significant increase in patients with more severe liver disease (**Fig.3q-r**).

**Figure 3.**
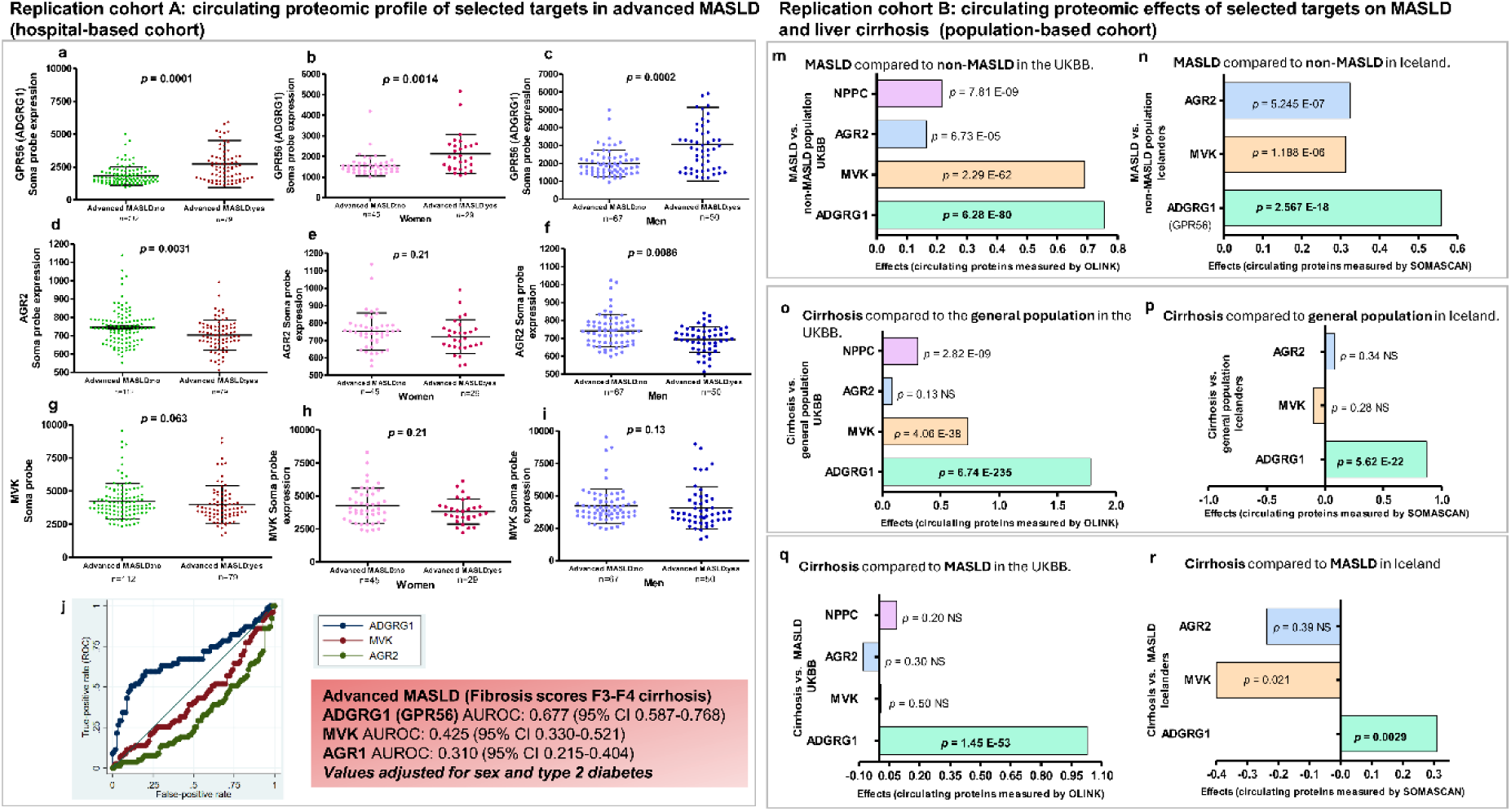
Circulating proteomic profile of selected targets in two different replication cohorts. **Replication cohort A:** Chars show association for selected targets from the discovery phase, including ADGRG1, AGR2, and MVK. Metadata of 191 plasma samples processed for proteomics using the SomaScan v.4.0 platform and accessed from ref. [28]. Proteomic profile of ADGRG1 (**a,b,c**), AGR2 (**d,e,f**), and MVK (**g,h,i**) in patients with MASLD classified as advanced MASLD (fibrosis stage F3-F4) versus no advanced-MASLD (fibrosis score F0–F2), and disaggregated by sex. *P* values are for Mann Whitney two-tailed test. **j.** AUROC curve statistics of the three biomarkers for detection of advanced MASLD adjusted for relevant covariates. **Replication Cohort B**: Charts show results of association effects between proteomic data of selected targets (ADGRG1, AGR2, MVK, and NPPC) and MASLD or cirrhosis compared to healthy general population in two different population-based studies accessed from ref. [32]. Association effects for circulating proteomic profile with MASLD diagnosis (*n*=610) compared to non-MASLD population (*n*=38018) in the UKBB and measured by OLINK platform (**m**); association effects with MASLD diagnosis (*n*=181) compared with non-MASLD population (*n*=20619) in the Icelanders sample and measured by SomaScan platform (**n**). Association of protein levels with cirrhosis diagnosis (*n*=262) in comparison with the general population (*n*=38018) in the UKBB and measured by OLINK platform (**o**); association of proteins with cirrhosis diagnosis (*n*=73) in comparison with the general population (*n*=20619) in the Icelanders sample and measured by SomaScan platform (**p**). Association of proteins with cirrhosis diagnosis (*n*=262) in comparison with MASLD (*n*=610) in the UKBB and measured by OLINK platform (**q**); association of proteins with cirrhosis diagnosis (*n*=73) in comparison with MASLD (*n*=181) in the Icelanders sample and measured by SomaScan platform (**r**). *P* values for association are two-tailed, unadjusted for multiple comparisons and are from a linear regression model with age and sex as covariates.

### Validation of data on incident outcomes and mortality of the selected target

After adjusting for age, sex, BMI, alcohol consumption, smoking status, education status, and social deprivation, five associations of ADGRG1 circulating levels with major outcomes, including death, remained significant (**Fig.4a-f**). The strongest and most significant effects were observed for ADGRG1 plasma levels and the risk of end-stage liver disease and T2D (**Fig.4b-c**), with hazard ratios (HR) of 2.94 and 1.72, respectively. ADGRG1 was also associated with a small (1.26 HR) yet clinically and statistically significant risk of death over 16-years of follow up (**Fig.4d**). ADGRG1 levels were associated with a relatively modest effect on the risk of chronic obstructive pulmonary disease (COPD) (**Fig.4e**) and ischaemic heart disease (**Fig.4f**).

**Figure 4.**
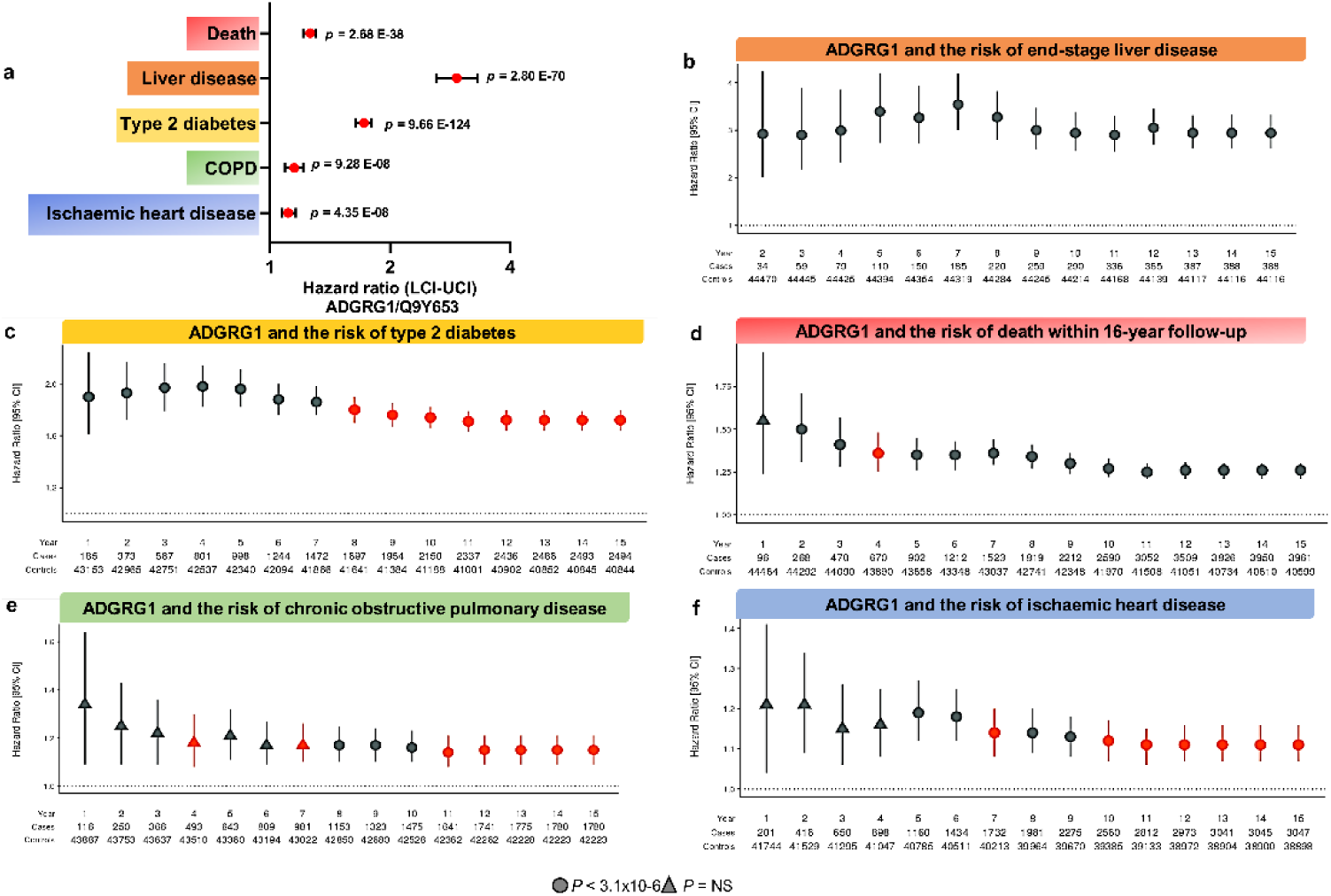
UKBB incident common diseases charts and mortality data associated with ADGRG1 levels. **a.** Cox proportional-hazard model associations between ADGRG1 and five significant incident outcomes in the UKBB proteomics study. **b-f**. Significantly associated conditions and death with ADGRG1 plasma protein levels. *P* values are fully adjusted for age, sex and six lifestyle covariates. HR: hazard ratio. LCI: lower confidence interval. UCI: upper confidence interval. Circles indicate associations with P < 3.1×10-6 (Bonferroni threshold), whereas triangles indicate no association. Violations of the Cox PH assumption (Schoenfeld P < 0.05) at the local protein level are shown in red. Charts were done using the Shiny app available at: https://protein-disease-ukb.optima-health.technology.

### Protein-phenotype associations in the UKBB population

Protein-phenotype associations in the UKBB population showed significant associations between circulating levels of ADGRG1 and liver phenotypes, particularly liver failure and cirrhosis, including oesophageal varices, as well as quantitative laboratory traits such as gamma-glutamyl transferase and alkaline phosphatase (**Table S3**). There were also associations with major CVD, such as acute myocardial infarction, chronic ischaemic heart disease, heart failure and arterial hypertension; neurological diseases, such as cerebral infarction and stroke; kidney diseases, such as acute and chronic renal failure; and respiratory diseases, such as emphysema and other COPD (**Table S3**). The effects were consistent across all ancestry groups.

## Tissue origin of circulating biomarkers of organ damage

ADGRG1 protein expression is weak in isolated hepatocytes in MASL (0.17±0.25), whereas the immunostaining pattern in samples with MASH-fibrosis is strong (2.25±0.98) with a prominent epithelial expression profile in hepatocytes located mainly adjacent to portal tracts, periseptal areas, and areas of inflammatory infiltrate, particularly in nuclei (**Fig.5a**); *p* =0.0043 Mann-Whitney U Test. Although there was an increased number of positive cells in biopsies from patients with MASH fibrosis compared to MASL, quantification of immunostaining did not show significant differences for MVK, NPPC and AGR2; **Fig.S4.**

### Liver gene expression signature of validated circulating proteins

The analysis of liver bulk RNA-seq data showed that *ADGRG1* expression is significantly and consistently upregulated in the liver of patients with advanced fibrosis in two independent samples of patients (**Fig.5b,c**).

**Figure 5.**
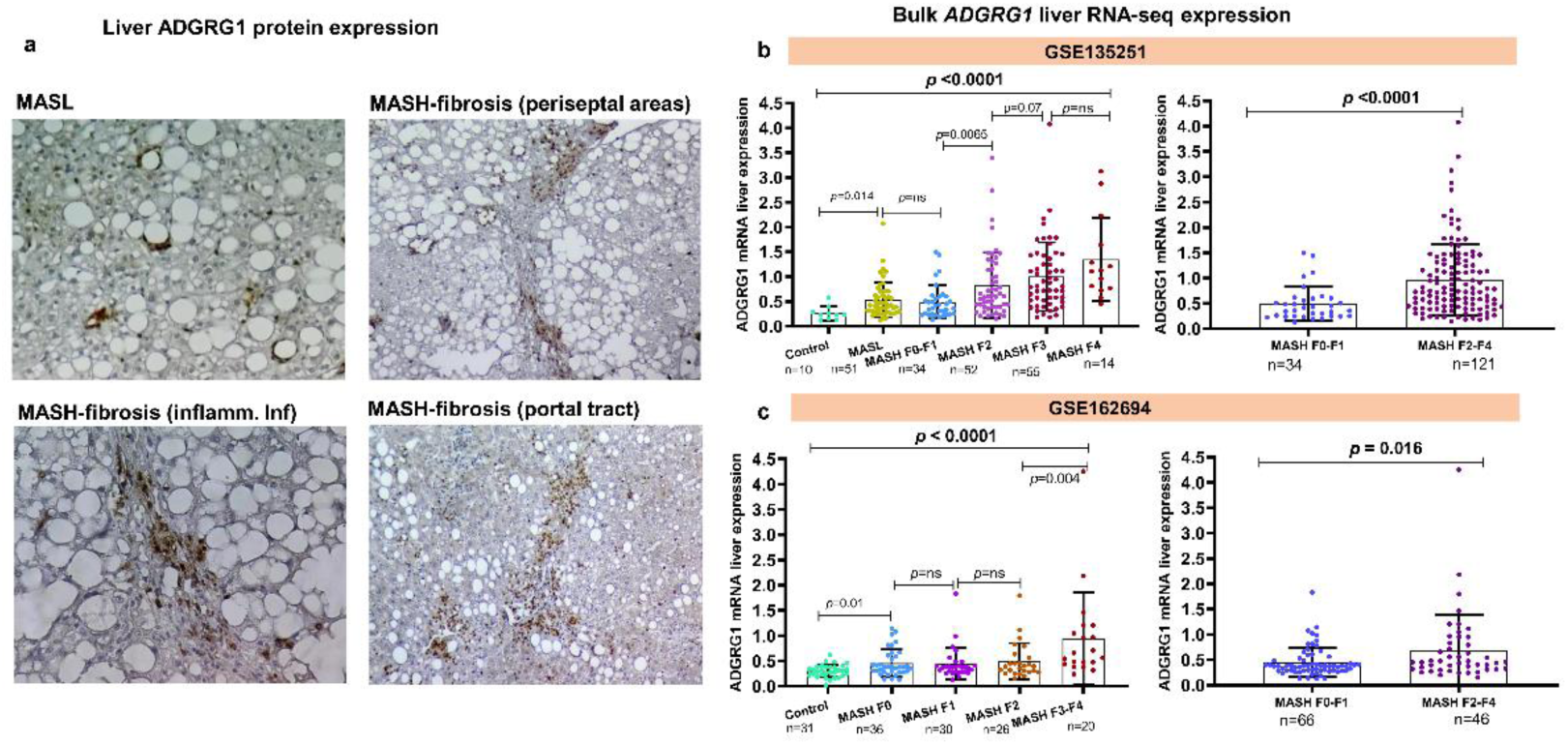
ADGRG1 protein and gene expression in the liver mirror the circulating profile. Representative immunohistochemistry images for ADGRG1 in human liver biopsies. Arrows indicate the positive immunostaining in periseptal and portal areas, and inflammatory infiltrate (inflamm. Infiltrate) (**a**). Original magnification: ×200. Bulk liver ADGRG1 expression in MASH-fibrosis (**b,c**). Bulk liver transcriptome data accessed from the NCBI GEO repository (GSE135251 n=216 and GSE162694 n=143). Mann–Whitney-*U* test (two-tailed). *P*-values as indicated in the figure, ns, not significant.

### *ADGRG1* gene expression at the individual cell-level resolution in the healthy liver

Results indicates that in the absence of a liver injury stimulus, *ADGRG1* expression is very low and restricted to immune-related cells (**Fig.S5a-c**). Uniform manifold approximation and projection (UMAP) visualization of profiled cells showed *ADGRG1* is predominantly expressed in natural killer (NK) cells. Examination of liver scRNA-seq lymphoid cell transcriptional profiles confirmed that *ADGRG1* is predominantly expressed in resident and circulating NK cells (**Fig.S5d-f**), albeit at very low levels. The ADGRG1 protein scRNA-seq signatures were mapped onto the Visium zoning data, confirming the zonation observed in the portal and periportal zones in the presence of steatosis, as well as the low levels of expression observed in the liver tissue in the non-steatotic tissue (**Fig.S5g-i**).

### *ADGRG1* cell type-specific transcriptional changes in a HLO model of steatohepatitis

To dissect cell type-specific changes in liver *ADGRG1* expression under steatogenic conditions, we looked at the effects of oleic acid (OA), a monounsaturated fatty acid, and palmitic acid (PA), a saturated fatty acid. Challenge with OA was associated with a significant increase in *ADGRG1* levels in all cells in the model except smooth muscle cells, fibroblasts and fetal hepatocytes (**Fig.6a,b**). Challenge with PA was associated with a significant increase in *ADGRG1* levels, particularly in hepatoblasts, fetal hepatocytes, cholangiocytes and adult hepatocytes (**Fig.6c,d**). Most importantly, significant changes in *ADGRG1* expression in hepatic stellate cells, smooth muscle cells, adult hepatocytes and ductal cells were associated with TGFβ1-mediated injury (**Fig.6e,f**), which recapitulated a liver fibrotic phenotype in the model [36]. Violin plots showing the number of single-cells and ADGRG1 transcripts for each condition are depicted in **Fig.S6**. Enriched gene ontology (GO) biological pathways associated with liver *ADGRG1* expression in response to the inducing stimulus are shown in **Tables S4-S6**.

**Figure 6:**
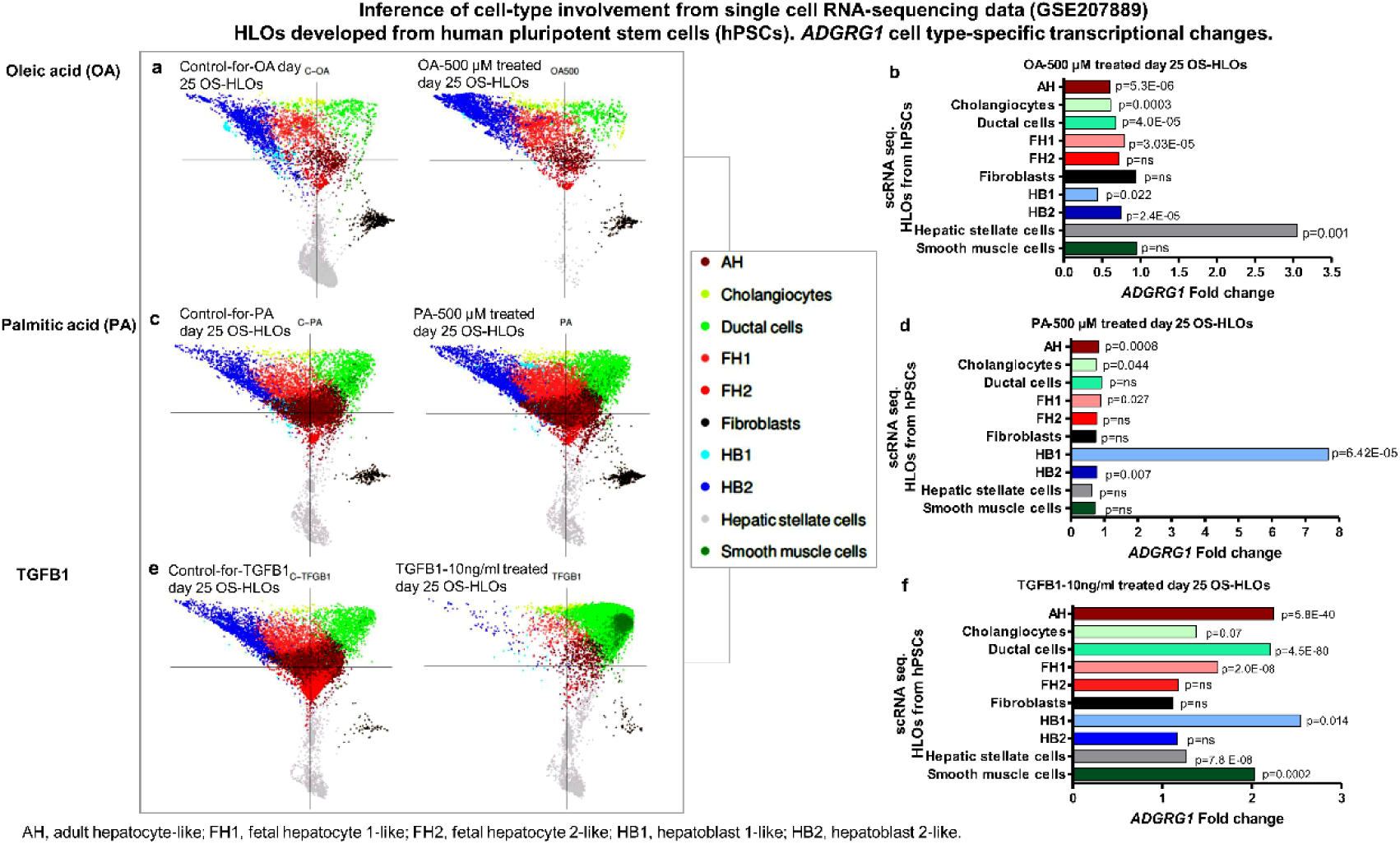
ADGRG1 is expressed in the cell of the fibrotic niche. **a,c,e**. ForceAtlas2 plots of cells from OS-HLOs treated for 4 days with OA (500 μM), PA (500 μM), or TGF-β1 (10 ng/ml) for their potential to model inflammation and fibrosis, and their respective controls (C-OA, C-PA, C-TGFBA). **b**,**d,f.** *ADGRG1* gene expression changes at single cell resolution (10× scRNA-seq transcriptomic analysis) in the three conditions. Mann–Whitney-*U* test (two-tailed). *P*-values as indicated in the figure, ns, not significant. OS-HLOs: human liver organoids cultured on an orbital shaker.

The top significantly enriched terms for *ADGRG1* in OA-treated cells correlate with GO pathways associated with vasculogenesis, integrin-mediated cell adhesion, cardiac epithelial to mesenchymal transition, SMAD protein phosphorylation, endodermal cell differentiation and epithelial cell apoptotic process (**Table S4**). In PA-treated cells, *ADGRG1* significantly correlated with GO pathways associated with integrin-mediated cell adhesion and signalling pathways, foam cell differentiation, vascular endothelial growth factor production and vasculogenesis, myotube differentiation and interleukin-8 production (**Table S5**). In fibrotic injury model cells stimulated with TGF-B1, *ADGRG1* expression significantly correlated with GO pathways associated with synaptic signalling, epidermal, keratinocyte, and stem cell differentiation, substrate adhesion-dependent cell spreading, myocardial cell action potential involved in contraction and myocardial cell contraction, ERK1 and ERK2 cascade, and myeloid leukocyte differentiation, among many other cancer-related pathways (see full list in **Table S6**).

Predicted interactions in the liver tissue and cell context (**Fig.S7a-b**) in which the ADGRG1 protein functions were explored using the HumanBase resource. Analysis of ADGRG1 expression in hepatocytes shows robust scores for interaction with FGFR3, CYP2C9, PKP3, SFRP1, NOL3, and APOA1 among the top scores (**Fig.S7b**). Investigation of cell type involvement in the HLOs model showed that under stimulation with OA and PA, *FGFR3* and *APOA1* parallel the gene expression changes of *ADGRG1* in adult hepatocytes, but not in other liver cells (**Fig.S7c-d**). Upon fibrotic injury, *FGFR3* gene expression levels paralleled *ADGRG1* gene expression changes not only in adult hepatocytes but also in ductal and hepatic stellate cells (**Fig.S7e**), indicating that *FGFR3* also shifts its expression profile towards a profibrotic phenotype.

## DISCUSSION

The involvement of MASLD in multisystem complications has been consistently demonstrated in clinical epidemiological studies [24]. However, the major challenge is the lack of clinically actionable biomarkers to monitor and assess multi-organ damage in patients with MASLD, in part due to insufficient knowledge of its precise molecular mechanisms.

Here, we used a high-throughput targeted proteomic approach that enables the detection of signatures of pathophysiological relevance in organ damage in small amounts of biological samples. We demonstrated that the proteins involved in MASLD-related organ damage, including ADGRG1, are primarily deregulated in patients at-risk of progressive disease and having more severe histological stages.

As well as addressing a clinical need, this finding has several implications. First, it is notable that when the replication cohorts were examined, the circulating levels of ADGRG1 exhibited a similar direction of change and effects across both the disease stages and the proteomic platforms, which may diverge for most of the proteins [38]. The replication includes the trajectory of ADGRG1 levels in patients with varying degrees of liver fibrosis, which have similar increased directionality from mild fibrosis to cirrhosis. Consistent with this observation, Govere et al. found that ADGRG1 was among the top ranked circulating proteins that significantly correlated with transcriptomic expression, found only in patients with significant fibrosis among 4,584 protein probes [28]. This is important as it enables the stratification of patients into MASLD-molecular subtypes, which are associated with an increased risk of extrahepatic complications. Concurrently, cross-platform validation demonstrating a robust correlation between protein effects and a notable association with liver phenotypes underscores the precision and/or utility of the identified analyte [38].

Second, validation data suggest that ADGRG1 levels may serve as a predictor of mortality, with a hazard ratio of 1.26, and may indicate an increased risk of complications associated with liver disease, such as cirrhosis, respiratory disease, T2D and CVD, independent of age, sex or lifestyle covariates. Collectively, ADGRG1 may be used as a prognostic biomarker of multisystemic damage in advanced MASLD.

Third, using deconvolution strategies and exploration of potential origin of the involved targets we found that the affected liver may be responsible to produce organ-damage related proteins. Specifically, we found that ADGRG1 gene and protein expression were significantly upregulated not only in the circulation but also in the liver of patients with severe MASLD. This makes our results both plausible and robust, as at least on the bulk level, protein levels are largely determined by transcript concentrations [39]. It also implies that damage to other organs in the body can be prevented by regressing the fibrotic lesions, which emphasises the necessity of expediting the approval of MASH-drugs for the treatment of patients with moderate to advanced fibrosis [40].

Last, but not least, our results offer insights into the disease mechanisms that lead to organ damage in MASLD. ADGRG1, which is scarcely expressed in the healthy liver with low levels of gene expression in non-parenchymal cells such as NK-cells, demonstrated a pattern consistent with impaired protein homeostasis in MASH. Protein expression levels were significantly upregulated in MASH-fibrosis, with a notable increase observed in periportal hepatocytes and septal areas. Single-cell transcriptomic modelling revealed that in addition to hepatocytes and ductal cells, cells of the fibrotic niche are involved in the expression of ADGRG1. The GO annotations related to ADGRG1 in the model are not only associated with epithelial cell differentiation, but also pathways linked to cardiac muscle cell contraction and conduction, as well as synaptic signalling.

ADGRG1 (GPR56) is a member of the G protein-coupled receptor family (GPCRs) that plays a key role in cell adhesion and cell-cell interactions, signal transduction, cancer progression, and in multipotent cell identity [41]. It has several binding partners. Collagen III and tissue transglutaminase 2, a large calcium-dependent enzyme involved in cytoskeletal regulation, both interact with ADGRG1 [42]. Once released into the circulation, ADGRG1 can act as a mediator of cross-tissue communication.

The assessment of specific protein-hepatocyte interactions revealed biological associations between ADGRG1 and other proteins in the network, including FGFR3 that may explain the involvement of this protein in extrahepatic complications, including cancer and systemic fibrogenesis. This is of interest because systemic fibrogenesis in the lungs, kidneys, liver and cardiovascular system, as well as impaired tissue renewal and remodelling, are major causes of morbidity and mortality [43] [44].

This study has limitations. First, the underlying molecular processes and disease mechanisms of extrahepatic complications in MASLD could be explained by a myriad of deregulated proteins and the co-occurrence of multimorbidity, socioeconomic disparities, genetic predisposition and environmental factors that may independently influence the burden of chronic complex diseases. Therefore, the heterogeneity of MASLD [45] [46] makes it difficult to provide answers to all causal and pathological mechanisms and associations with extrahepatic diseases. Second, although several strategies were employed for replication and validation of the selected target in diverse study designs, and large and diverse populations, further research is needed to assess the generalisability of the findings in more ethnically and geographically diverse populations, as genetic variation, among many other factors, could be a source of variation in protein levels [38].

In conclusion, a comprehensive systemic assessment is necessary to evaluate patients with MASLD at risk of extrahepatic complications. Surrogate markers of inflammation that improve risk scores for predicting the long-term prognosis [47] and imaging techniques and non-invasive biomarkers of fibrosis [48] [7] [49] are not designed to track the extrahepatic complications associated with MASLD at the molecular level. The strength of our study is the use of proteomics to identify a mechanistic GPCR-biomarker associated with multisystem damage in patients with progressive MASLD. This finding is both plausible and biologically sound, as deregulation of GPCRs has been identified as a contributing factor in numerous major complex human diseases, including cancer, cardiovascular disease, diabetes, neurological disorders, inflammation, and obesity [50]. Therefore, our study suggests that ADGRG1 can be used for MASLD-subtype characterisation and personalised risk assessment of extrahepatic complications, as well as for holistic prevention, diagnosis and monitoring of systemic organ damage. The implementation of the identified biomarker can also be adapted to provide rapid and cost-effective point-of-care results. The accessible cell surface locations that can be targeted by small-molecule drugs [51] make ADGRG1 a potential drug target for preventing organ damage in patients with MASLD.

## Abbreviations

ADGRG1: Adhesion G-Protein Coupled Receptor G1.

AGR2: Anterior Gradient 2, Protein Disulphide Isomerase Family Member.

AUROC: area under the receiver operating characteristic.

BMI: body mass index.

COPD: chronic obstructive pulmonary disease.

GEO: Gene Expression Omnibus.

GO: Gene Ontology.

GPCRs: G protein-coupled receptors

HR: hazard ratio.

HLOs: human liver organoids.

HCC: hepatocellular carcinoma.

MASLD: metabolic dysfunction-associated steatotic liver disease.

MASL: metabolic dysfunction-associated steatotic liver.

MASH: metabolic dysfunction-associated steatohepatitis.

MVK: Mevalonate Kinase.

NAS: NAFLD activity score–clinical research network.

NPPC: Natriuretic Peptide C

ScRNA-seq: single cell RNA sequencing.

OA: oleic acid.

OR: odds ratio

PA: palmitic acid.

PEA: Proximity Extension Assay.

T2D: type 2 diabetes.

TGFB1: Transforming Growth Factor Beta 1.

UKBB: U K Biobank.

UMAP: uniform manifold approximation and projection.

## Data Availability

The authors confirm that all relevant data generated in this study are included in the article and/or its supplementary information. Clinical data in this study can be found in Table 1 and Table S2. In accordance with the Ethics Committee of our institution and the Institutional Review Board of the GCBA regarding the protection of individual privacy, personal and sensitive data associated with phenotypic and proteomic data will only be available for controlled access and will be available from the corresponding author upon reasonable request. ADGRG1 protein-phenotype associations in the UKBB population stratified on ancestry can be found in Table S3. All RNA-seq data are publicly available from the Gene Expression Omnibus database (accession numbers (GSE135251, GSE162694) or Liver Cell Atlas (accession number GSE192742; Scripts Liver Atlas data: github.com/guilliottslab). Inference of cell-type involvement from single cell RNA-sequencing data are publicly available in GSE207889, and additional reanalysis of single cell expression can be found in Figure S7. Enriched gene ontology (GO) biological pathways associated with liver ADGRG1 expression in response to the inducing stimulus can be found in Tables S4-S6.

## Supplementary information titles and legends

**Figure S1:**
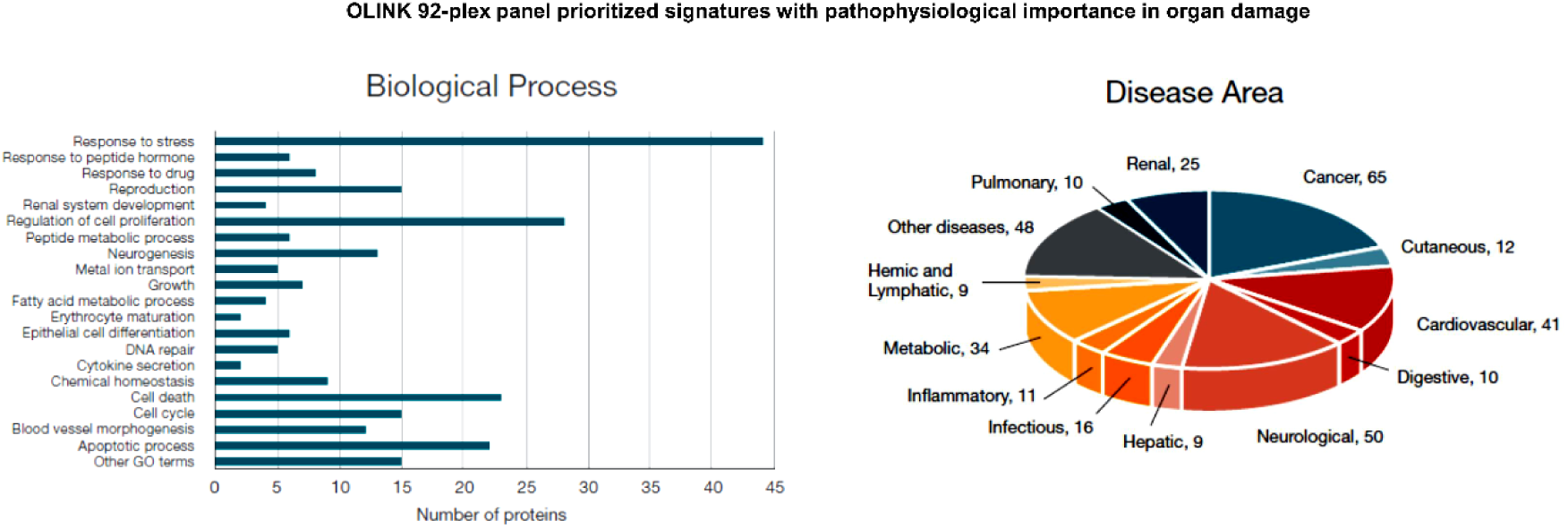
Organ damage proteomics panel. Charts show the list of 92 biomarkers included in the organ damage panel developed by OLINK are classified according to Biological Process and Disease Area (based on public-access bioinformatic databases, including Uniprot, Human Protein Atlas, Gene Ontology (GO) and DisGeNET.

**Figure S2.**
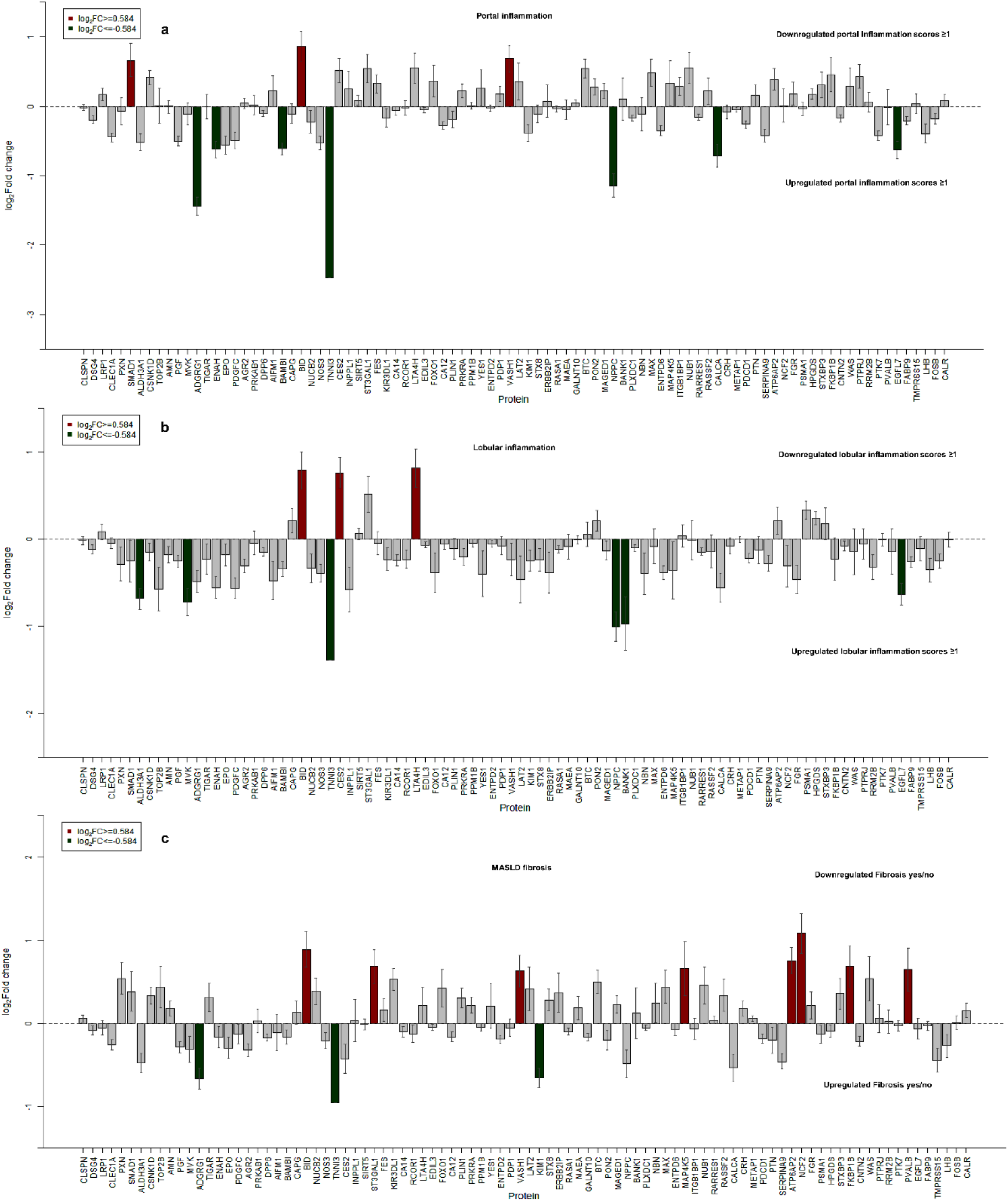
Discovery phase using a validated organ damage proteomic assay: analysis of histological traits. Log_2_(fold change) of differentially expressed proteins **a.** Portal inflammation: 0 (none)-1 (mild) versus 2 (moderate)-3 (marked); **b.** Lobular inflammation: 0-3 and comparing none versus presence based upon the number of foci per 20x fields); **c.** MASH-without fibrosis versus MASH-fibrosis.

**Figure S3:**
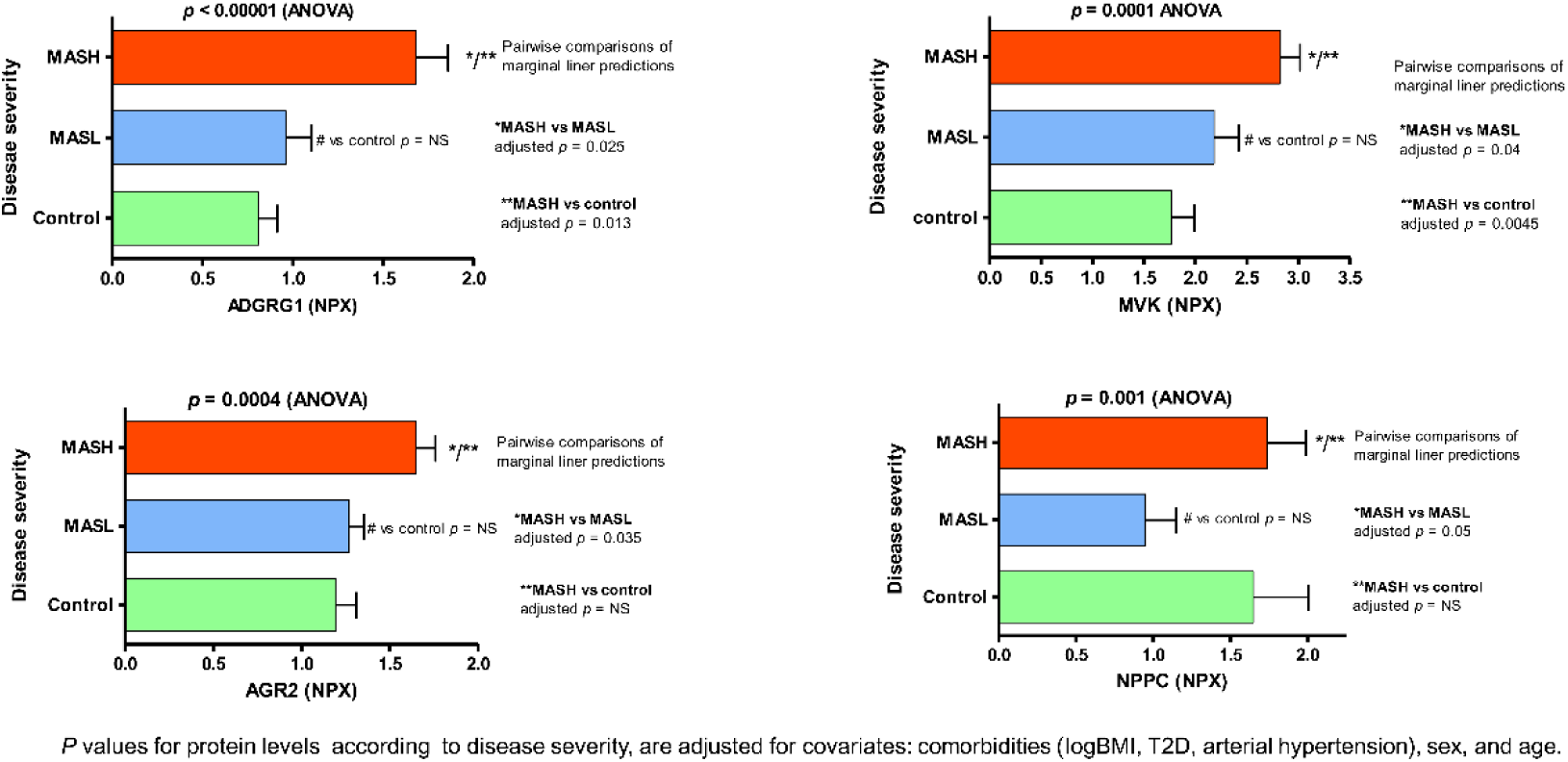
Selected targets deregulated in MASLD across the full histological spectrum.

**Figure S4:**
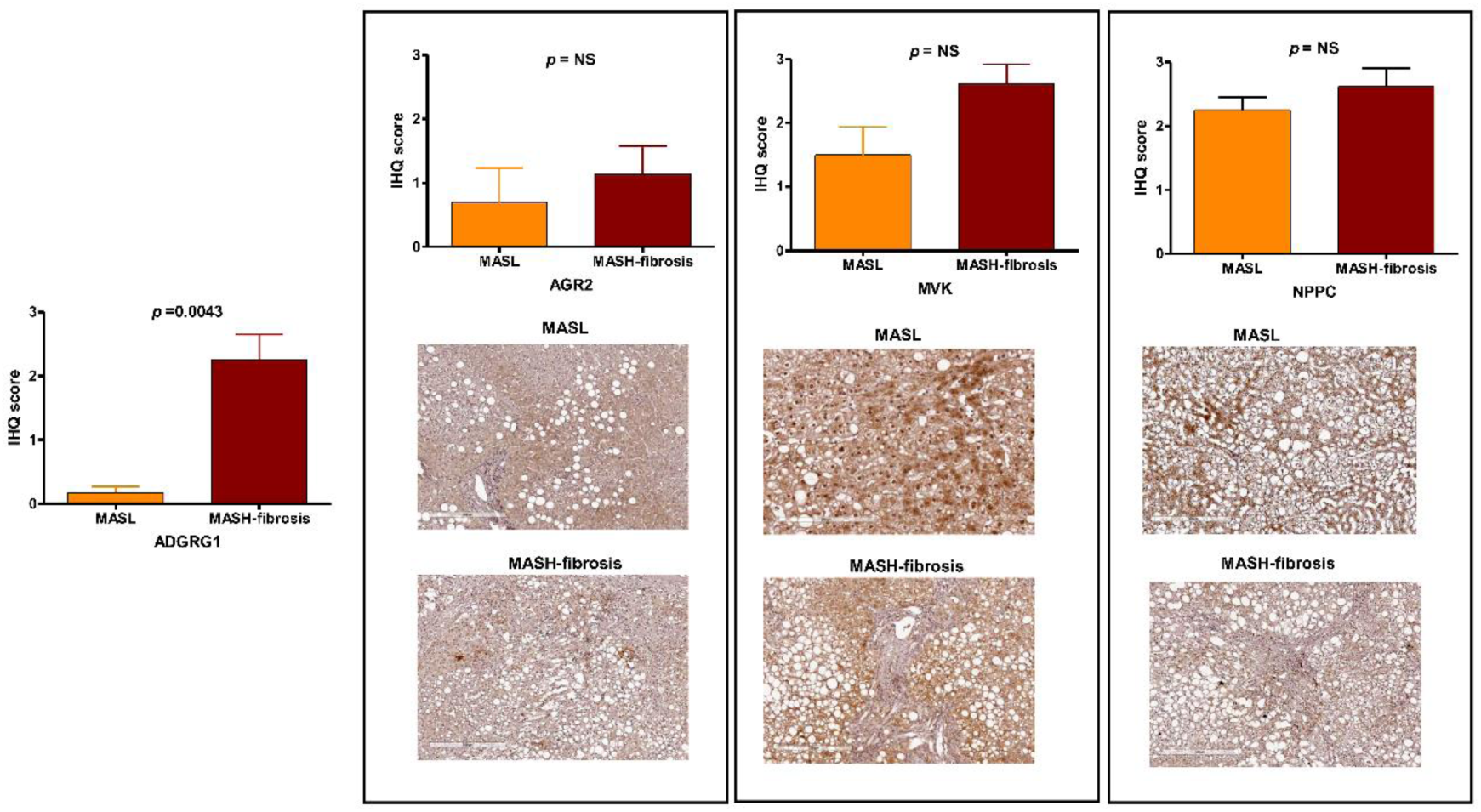
Immunohistochemical staining scores and representative immunohistochemistry images in human liver biopsies. Selected targets from the discovery phase (ADGRG1, AGR2, MVK, and NPPC) in 31 biopsies (12 specimens for each protein) from patients with MASLD across the severity spectrum (MASL and MASH-fibrosis).

**Figure S5:**
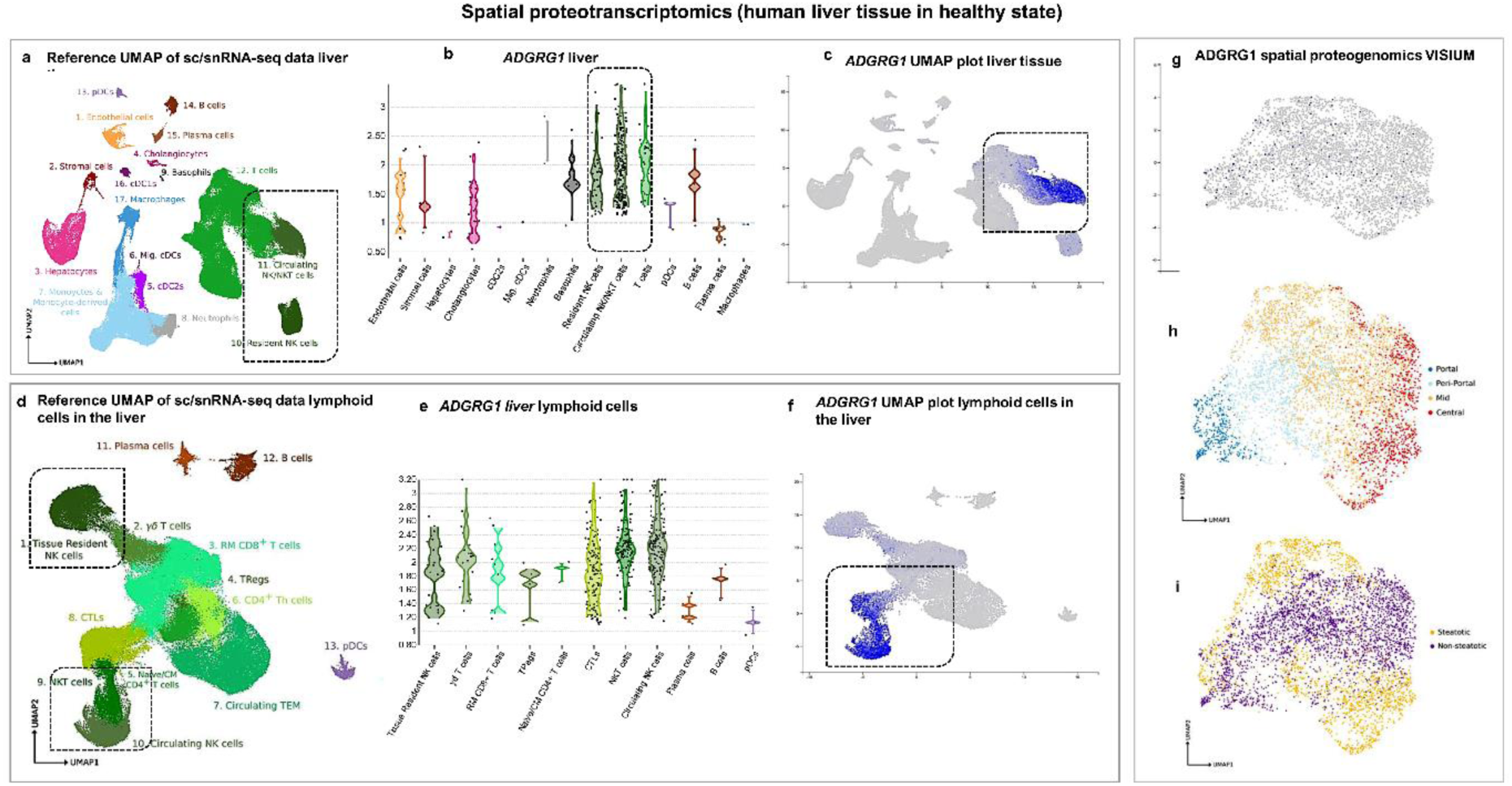
Spatial transcriptomics profile of *ADGRG1* in human liver tissue in healthy state. We interrogated the spatially resolved gene expression profile of *ADGRG1* in cellular niches of human liver in healthy state using the Liver Cell Atlas available https://www.livercellatlas.org/umap-humanAll.php. **a**. Reference UMAP visualization of sc-RNA seq data of liver cells based on healthy human tissue. **b**. *ADGRG1* transcript abundance in each cell type present in the heathy human liver. **c.** *ADGRG1* UMAP visualization of sc-RNA seq data of liver cells based on healthy human tissue. **d**. Reference UMAP of sc/snRNA-seq data lymphoid cells in the healthy liver. **e**. *ADGRG1* gene expression profile in liver lymphoid cells in healthy state. **f**. UMAP visualization of sc-RNA seq data for *ADGRG1*in lymphoid cells in the healthy liver. **g**. Singe cell proteogenomics: mapping of *ADGRG1* expression, Visium UMAP zonation and expression patterns in healthy liver. **h**-**i**. Reference mapping of Visium UMAP zonation patterns in steatotic and healthy liver modelled from the Liver Cell Atlas.

**Figure S6:**
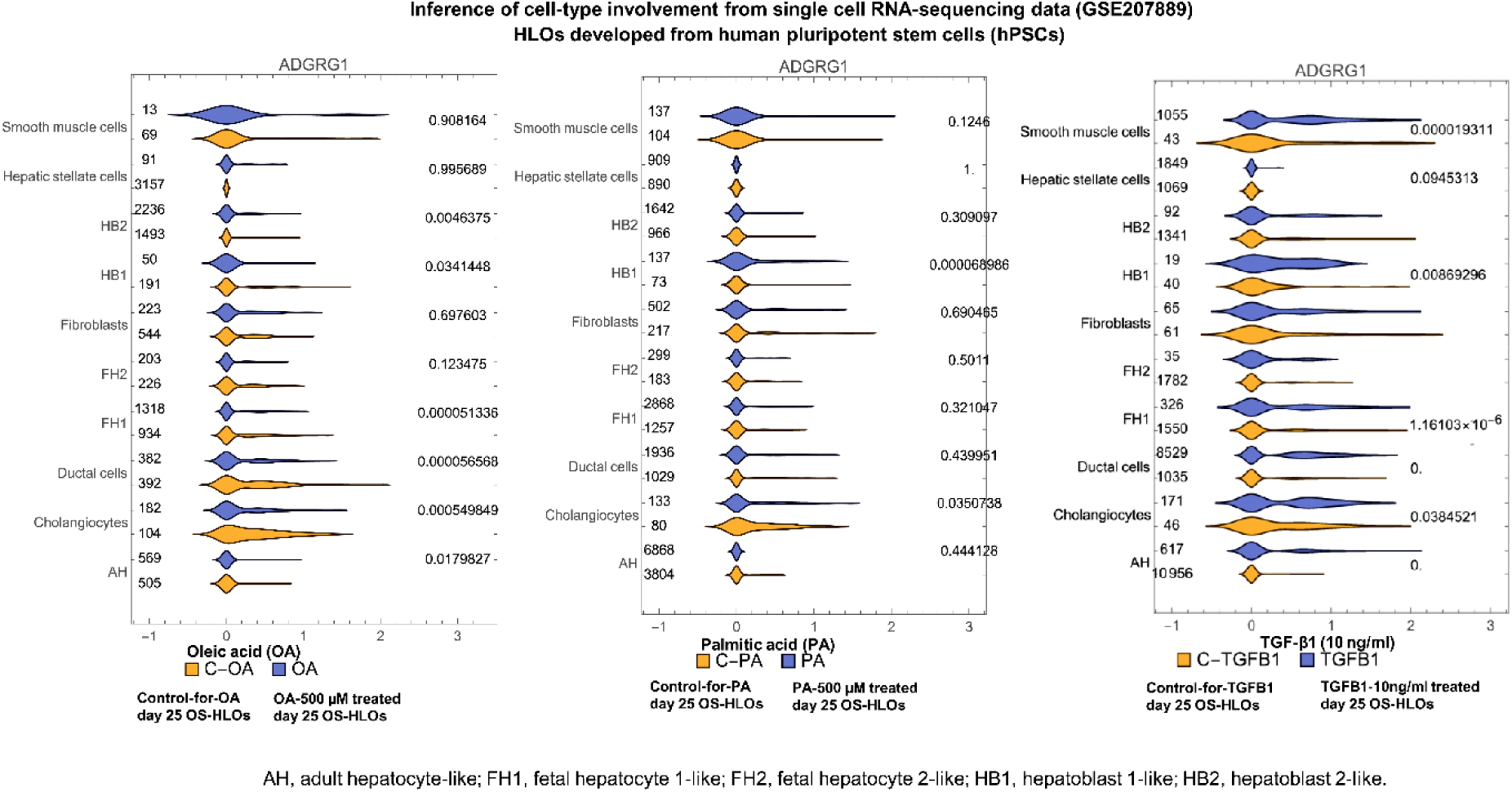
Inference of cell-type involvement from single cell RNA-sequencing data (GSE207889) violines.

**Figure S7:**
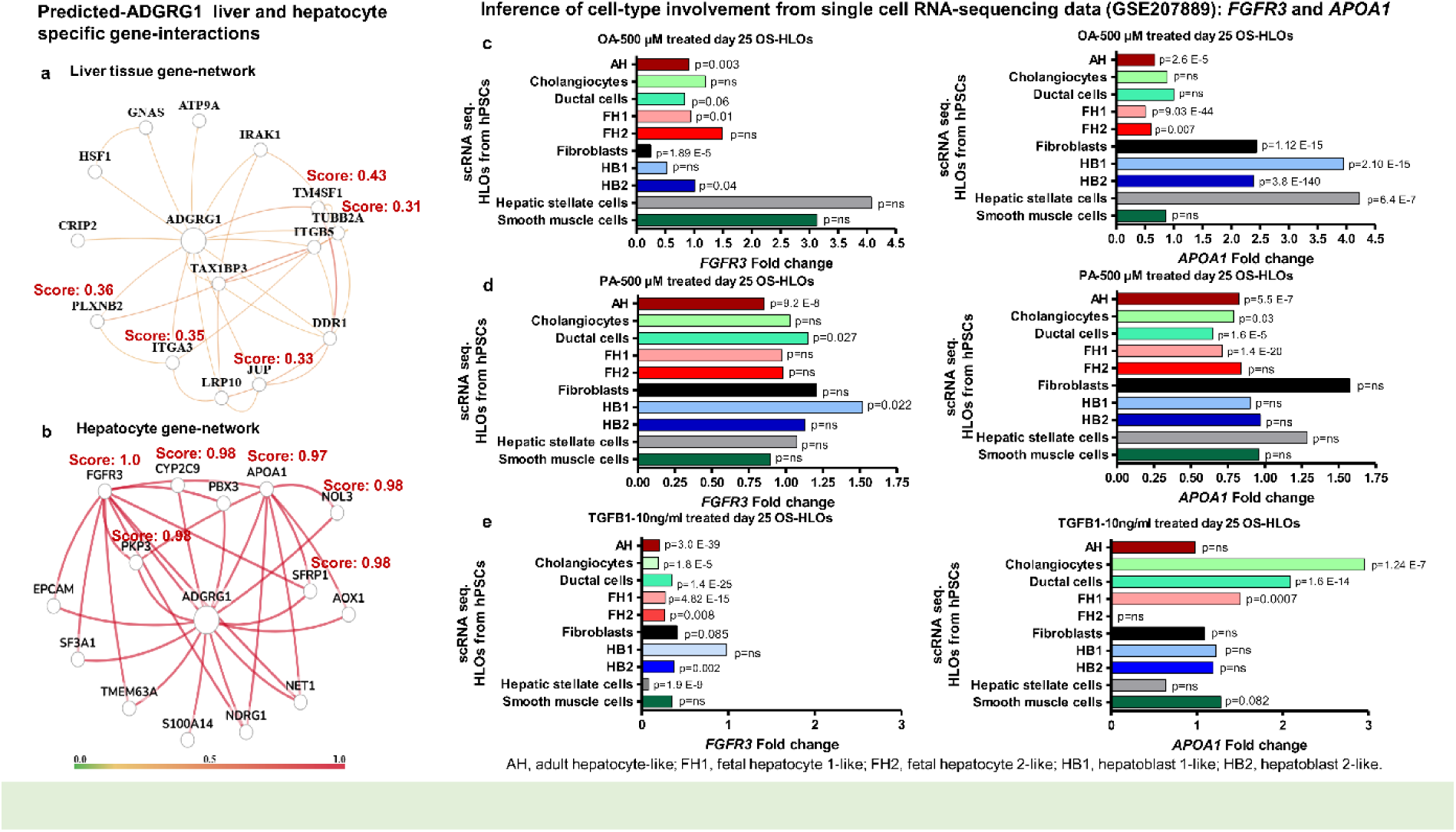
ADGRG1 tissue and liver cell-specific gene interactions. Predicted interactions of ADGRG1 in the liver tissue (**a**) and hepatocytes (**b**) were performed using genome-wide functional interaction networks for human tissues and cell types developed using a data-driven Bayesian methodology accessed from the HumanBase available at https://hb.flatironinstitute.org/. The predicted functional networks show edges based on co-expression, interactions, transforming factors binding, GSEA microRNA targets, and GSEA perturbations. GO: gene ontology pathways. **c-e**. *FGFR3* and *APOA1* cell type-specific transcriptional changes in a model of MASH based on human liver organoids developed from human pluripotent stem cells. Pre-processed scRNA-seq data were accessed from GSE207889; details of the model were accessed from ref. [36]. Cells from OS-HLOs treated for 4 days with OA (500 μM), PA (500 μM), or TGF-β1 (10 ng/ml) for their potential to model inflammation and fibrosis. *FGFR3* and *APOA1* gene expression changes at single cell resolution (10× scRNA-seq transcriptomic analysis) in the three conditions. Mann– Whitney-*U* test (two-tailed). *P*-values as indicated in the figure, ns, not significant. OS-HLOs: human liver organoids cultured on an orbital shaker.

## Supplementary Methods

### Discovery phase: Recruitment strategy, and clinical and histological data collection

Individuals in the discovery phase were divided into three groups: those without MASLD, who were regarded as controls (n=10) and those with histologically confirmed MASL (n =35) and MASH (n=43). A matched cohort study design was used to ensure that the non-MASLD liver samples had similar patient characteristics, including demographics and anthropometrics. Participants self-reported their sex, and there was no gender bias in the study design. Biological samples were chosen based on availability, quality, and integrity. The study included patients who had histopathologic evidence of fatty liver disease, specifically MASL or MASH, on a liver biopsy conducted during the study period and sample collection. The selection of biological samples for the current study was based on the availability of an adequate amount of high-quality serum to perform proteomic analysis. Exclusion criteria were applied, including secondary causes of steatosis (e.g., alcohol abuse, total parenteral nutrition, viral hepatitis, drug use), autoimmune liver disease, metabolic liver disease, Wilson’s disease, and alpha-1-antitrypsin deficiency. Non-MASLD serum samples were selected from patients without evidence of MASLD or metabolic syndrome, whose age and sex matched those of the patients with MASLD. These patients had near-normal liver histology in specimens obtained by liver biopsy. Only participants without fatty change on histological examination were included in the study for non-MASLD subjects. Biological samples were consecutively selected from participants with and without MASLD during the same study period and from the same patient population in Argentina. The matched patients shared the same demographic characteristics, including occupation, educational level, place of residence, and ethnicity. Arterial hypertension was defined as a systolic blood pressure of at least 140 mm Hg and/or a diastolic blood pressure of at least 90 mm Hg, determined by multiple examinations and averaging two or more blood pressure readings on two subsequent visits. Liver specimens were obtained during bariatric surgery or using a modified 1.4-mm-diameter Menghini needle (Hepafix, Braun, Germany) via liver biopsy under local anaesthesia with ultrasound guidance or outpatient. Samples from the left lobe were collected right after opening the abdomen. Each liver biopsy sample was immersed in 40 g/l formaldehyde (pH 7.4), embedded in paraffin, and stained with haematoxylin and eosin, Masson trichrome, and silver impregnation for reticular fibres. Each biopsy was at least 3 cm long and had a minimum of eight portal tracts.

Samples from patients with MASLD were histologically scored according to the widely used NASH-Clinical Research Network semiquantitative NASH Activity Score (NAS) [1] for lobular inflammation (0–3), ballooning (0–2) and steatosis (0–3) and categorised according to histopathological disease grade and stage into MASL and MASH with different fibrosis stages (F0–4). Portal inflammation was scored as 0 (none), 1 (mild) or 2 (more than mild) according to Kleiner et.al. [2]. Patients with MASLD at risk of disease progression were classified as having a NAS score more than or equal to 4 with fibrosis stage of 2 or more (*F* ≥ 2) [3] [4] [5], unless indicated otherwise. Clinically significant fibrosis was defined as having a fibrosis stage ≥2, advanced fibrosis as ≥stage 3, and cirrhosis as stage 4 [6].

### Replication phase: Recruitment strategy, and clinical and histological data collection

Patients in the replication phase A and B were recruited as explained elsewhere in ref. [3,7]. Samples from the replication cohort A were derived from the European NAFLD Registry (NCT04442334), and liver samples were centrally scored according to the semiquantitative NASH-CRN Scoring System by an expert liver pathologist [3]. Data reused and analysed in this study was extracted from publicly extended data accessed from ref. [3].

Patients in phase B of the replication study were selected from a population of approximately 500,000 individuals across the United Kingdom who participated in a large prospective cohort study. The participants were between 40 and 69 years old at the time of recruitment [7]. A subset of 38890 participants had proteomic data included in the analysis and accessed from ref. [7]. The disease status was determined using the ICD-10 code K76.0 for nonalcoholic fatty liver disease from electronic health records. The all-cause cirrhosis phenotype was identified based on cirrhosis- and fibrosis-related ICD-10 codes, including K70.2, K70.3, K70.4, K74.0, K74.1, K74.2, K74.6, K76.6, and KI85 [7]. The Icelandic deCODE genetics study is based on WGS data from 49,708 Icelanders [7]. Plasma samples were collected from Icelanders through two main projects: the Icelandic Cancer Project and various genetic programmes at deCODE genetics, Reykjavík, Iceland. The mean age of the participants was 55 years (s.d. = 17 years) and 57% were women [8]. All the considered reused data were within the Open Access Data Tier.

### Proteomics in the discovery phase

The proteomic Olink PEA platform was used to process 88 serum samples from the discovery phase (50 μl) in a 96-plex immunoassay for high throughput detection of protein biomarkers in body fluids [9]. This method is used in biomarker discovery with specific, highly targeted and validated panels associated with pathways, key biology process, or disorders of interest [10]. In this study, we used the biomarker to target organ damage. For each biomarker, a matched pair of antibodies linked to unique oligonucleotides binds to the respective protein target. When placed in close proximity, the stretch of nucleotides hybridises and the annealing product is then amplified by PCR and detected in multiplexed fashion in a high throughput fluidic chip system. Specificity and reproducibility of the assays are rigorously quality controlled [11].

Olink has a built-in quality control (QC) system using internal controls that enables full control over the technical performance of assays and samples. Three internal controls were spiked into every sample for each dilution and panel that are designed to monitor the quality of assay performance, as well as the quality of individual samples: immune control (incubation step), extension control (extension and pre-amplification step), and amplification control (amplification step). Each sample plate also included a control strip with the following controls: two sample controls used to estimate the precision (intra- and inter-CVs), three negative controls used to set the background levels for each protein assay and to calculate the level of detection, and three plate controls used to compensate for potential variation between runs and plates.

Olink Target 96 Organ Damage was analysed with 88 samples in one plate. Ct value was measured for 92 protein assay per sample and NPX value was calculated from Ct value using IPC normalized. Samples and protein assays were filtered by two strict thresholds (deviation from the control median NPX value, limit of detection, LOD). Following the application of the missing frequency, which depends on the limit of detection (LOD), 70 protein assays were retained.

QC analysis involved two stages: 1) run evaluation: calculation of standard deviations for incubation control and detection control should be within the pre-determined quality threshold (< 0.2), If > 1/6 of samples fail QC, there is high probability the entire run is unreliable. 2) Sample Evaluation: incubation control and detection control were assessed in each sample by calculation of deviation from the median value of each control respectively; a sample did not pass QC (flag / warning sample) if incubation control and/or detection control (corresponding to the specific sample) deviated +/- 0.3 NPX from the median value. The missing frequency plot indicates the limit of detection for all plates and the level of detectability. The frequency of samples that possess valid data was more than 75% of the total number of samples, the threshold required to declare the assay valid.

The final assay readout was expressed in NPX values, which is an arbitrary unit on a log2 scale in which a high value corresponds to high protein expression. NPX was calculated from Ct values and data pre-processing (normalization) was performed to minimize both intra- and inter-assay variation. Single plate analysis uses NPX values based on the median IPC for each protein; on the other hand, multi-plate normalization adjusts NPX values for each protein with intensity normalization so that the median of all samples is the same for all plates.

Detection of the digital PEA signal and sample analysis for Olink^®^ Explore were performed via NGS, using Illumina’s NovaSeq 6000 at Macrogen facility (Macrogen Seoul, South Korea). Data from the NovaSeq 6000 run was uploaded to the MyData cloud. After this, Olink MyData cloud software processed the data, performs QC-analysis, calculates NPX values and produces an Analysis Report. Fold change for each protein was visualized in bar plot, indicating the relative protein expression in each comparison. Each assay bars were colored in [Red (Up regulated) / Green (Down regulated)/Grey (Non - significant)] depending on the fold change. Further logistic and regression analysis with adjustment for confounder were realised in house as described in Statistical section (see below)

### Proteomics from replication cohorts

Samples from the replication A cohort were processed in the proteomic aptamer-based SomaScan Platform (SomaLogic) as reported elsewhere [3]. Access to protein expression values of selected protein targets was based on proteomics data available for the whole cohort (*n*=191) and the whole proteome, including data on sex/gender, T2D and advanced fibrosis, as well as SomaID and UniProt identifiers.

Samples from the replication B cohort were processed using the Olink Explore 1536 platform as part of the UKB-Pharma Proteomics Project, including measurement of the levels of 1,472 proteins. This sample included a subset of 38890 individuals in the UKBB study (MASLD *n*=610; cirrhosis *n*=262) [7]. In addition, samples from the Icelanders population (total sample *n*=20873; MASLD *n*=181; cirrhosis *n*=73) were measured with the SomaScan v.4 assay (Soma-Logic, Inc.) containing 4,907 aptamers, providing a measurement of relative binding of the plasma sample to each of the aptamers in relative fluorescence units (r.f.u.). Access to proteomics data (effects and p values) for associations of proteins with the selected diseases, as measured by either SomaScan or OLINK, was based on logistic regression with age and sex as covariates; however, two-tailed *p*-values not adjusted for multiple testing. Proteomics data was available for use and sharing in the source data from ref. [3] and ref [7].

### Validation of data on mortality and ADGRG1 plasma expression over 16 years and incident of 23-common diseases

Validation of longitudinal relationship between incident disease and mortality included data from the UKBB (n=47,600) using 1474 Olink protein analytes and accessed from ref. [12]. Analysis of ADGRG1 expression data in the general population across ancestry groups and its association with various diseases was based on proteomics data extracted from results for fair reuse and accessed from ref. [8].

Information on the incident of 23 age-related diseases and mortality recorded over 16 years of electronic health records (first occurrence) in the UK Biobank and or death registry (https://biobank.ctsu.ox.ac.uk/crystal/label.cgi?id=100093) and their significant association with the plasma proteome was retrieved from the publicly available information in Supplementary tables and datasets generated by Gadd et al. [12].

We used statistics (COX-proportions for hazard ratio HR, lower and upper confidence interval (CI), and *p* values) for circulating ADGRG1 levels, adjusted for age at baseline, and associations with mortality and incident of 23 outcomes that met the Bonferroni threshold of P < 3.1×10-6 accessed from ref. [12]. Models were adjusted for: age, sex and six lifestyle factors (BMI, smoking status, alcohol consumption, education status, physical activity and social deprivation) [12]. Quantification of protein levels was performed at Olink Analysis using Olink Proximity Extension Assay technology using four 384-plex panels (cardiometabolic, neurological, inflammatory and oncology) that targeted 1,463 unique proteins [12]. The results from the sensitivity analyses were visualised for every association tested in a Shiny app at: https://protein-disease-ukb.optima-health.technology. [12]. This resource allows graphing the number of cases over the 16 years of follow-up is shown for each characteristic, with the number of cases by year of follow-up plotted cumulatively, including fully adjusted *p* values for association. Complete list of incidents outcomes and coding index are as follows: cystitis (N30); multiple sclerosis (G35); brain/CNS cancer (C71,C72); schizophrenia (F20); systemic lupus erythematosus (M32); endometriosis (N80); vascular dementia (F01); amyotrophic lateral sclerosis (sourced from Motor Neurone Disease G12); inflammatory bowel disease (K51, K50); major depression (F33); gynaecological cancers (C51,C52,C53,C54,C55,C56,C57,C58); Alzheimer’s dementia (F00, G30); lung cancer (C34); rheumatoid arthritis (M05,M06); Parkinson’s disease (G20); colorectal cancer (C18,C20,C21); liver disease (K70,K71,K72,K73,K74); ischaemic stroke (I63); breast cancer (C50); prostate cancer (C61); chronic obstructive pulmonary disease (J40,J41,J42,J43,J44); type 2 diabetes (E11); ischaemic heart disease (I20,I21,I22,I23,I24,I25); death (Death registry).

All models adjusted for age at baseline. Proteomics data was extracted from data of the whole proteome and for 35,232 tested conditions/ diseases, from which there were 3201 associations involving 1,209 protein analytes and only 23 outcomes that met the Bonferroni threshold of *P* < 3.1×10-6 in the adjusted lifestyle-adjusted model. Data were accessed from ref. [12].

### Protein-phenotype associations in the general population UKBB across ancestry groups

The estimated association of protein levels with quantitative traits was done using linear regression. All analyses were adjusted for the sex and age of the individual at the time of plasma collection, and in addition, quantitative measures were inverse normal transformed. Associations are stratified by ancestry group as individuals with British or Irish, South Asian and African ancestry. Results are adjusted for multiple testing of protein–phenotype associations using Bonferroni adjustment for the number of assays on each platform (*P* < 1.0 × 10^−5^ for 4,907 assays on SomaScan and *P* < 1.7 × 10^−5^ for 2,941 assays on Olink). The description of main conditions/phenotypes is based on health care records [8]

### Liver protein expression of selected targets

Validation of hepatic protein expression of the selected biomarkers was performed by immunohistochemical staining of 12 cases per each protein of formalin-fixed paraffin-embedded liver tissue for each biomarker (ADGRG1, AGR2, MVK and NPPC) from the discovery cohort based on biospecimens availability. Liver protein expression of the identified targets involved 12 biologically independent samples of both sexes; MASL *n*=6 and MASH-fibrosis *n*=6. In total, samples of 31 different subjects were used. Four-micrometer sections were mounted onto silane-coated glass slides to ensure section adhesion through subsequent staining procedures. Briefly, sections were deparaffinized, rehydrated, washed in phosphate buffer solution (PBS), and treated with 3 % H_2_O_2_ in PBS for 20 min at room temperature to block endogenous peroxidase. Following heat-induced epitope retrieval in 0.1M citrate buffer at pH 6.0 for 20 min, the slides were incubated with a *specific antibody for ADGRG1 (GPR56) (MBS9604267) at a concentration of 1:100, MVK (MBS2526272) at a concentration of 1:100, AGR2 (MBS9209230) at a concentration of 1:200, and NPPC (MBS2517245) at a concentration of 1:150 (MyBioSource Inc, San Diego CA, United States)*.

Immunostaining was performed using the VECTASTAIN® Universal Quick HRP Kit (Peroxidase), R.T.U. (Ready-to-Use) (Vector Lab. CA, USA) detection system (PK-7800). Subsequently, slides were immersed in a 0.05% 3,3’-diaminobenzidine solution in 0.1 M Tris buffer, pH 7.2, containing 0.01 % H_2_O_2_. After a brown colour developed, slides were removed, and the reaction was stopped by immersion in PBS. Negative controls were carried out by omission of primary Ab. I*mmunostaining* was evaluated in a blinded fashion regarding any of the histological and clinical characteristics of the patients. Sections were counter-stained with Harris hematoxylin and examined by light microscopy in a blinded fashion (H-3401-500. Vector Lab. CA, USA).

The extent of staining was scored according to its amount and intensity by a 4-point scoring system as follows: 0 = no staining, 1 = positive staining in less than 20% of cells and /or tissue area of the portal tracts, 2 = 21-50 % of cells and /or tissue area of the portal tracts, and 3 = positive staining in more than 50% of cells and /or tissue area of the portal tracts. Microscopic evaluation was performed using a LEICA DM 2000 (Leica, Germany) trinocular microscope equipped with a high-definition *camera (Leica* MC190 *HD)*; all images were recorded using the Leica Application Suite (LAS) software.

### Bulk liver transcriptomics data

We used bulk liver transcriptome data from the GEO repository (GSE135251 and GSE162694). The data were further analysed using GEO2R. GSE135251 has expression profiling by high throughput sequencing of 216 snap-frozen biopsies (control samples *n*=10, MASL *n*=51, MASH F0-F1 *n*=34, MASH F2 *n*=52, MASH F3 *n*=55, MASH F4 *n*=14) that were processed for RNA sequencing on the Illumina NextSeq 500 system from ref. [13]. Groups were defined for the analysis as MASH with significant fibrosis (F2-F4 *n*=121) and MASH without significant fibrosis (F0-F1 *n*=34). GSE162694 dataset included 143 samples of patients with MASH across the full spectrum of fibrosis stage (F0 *n*=36, F1 *n*=30, F2 *n*=26, F3 *n*=8, F4 *n*=12) and control samples (*n*=31), which were processed for RNA sequencing on the Illumina HiSeq 3000 from ref. [14]. Groups were defined for the analysis as MASH with significant (F2-F4 *n*=46) and MASH without significant fibrosis (F0-F1 *n*=66).

### Spatial liver proteogenomic atlas

We interrogated the liver atlas dataset available at https://www.livercellatlas.org/umap-humanAll.php for gene and protein expression profiling of ADGRG1. This resource combines single-cell CITE-seq, single-nuclei sequencing, spatial transcriptomics, and spatial proteomics of the healthy human liver [15]. Human data is available at GSE192742. The Scripts Liver Atlas data is available at github.com/guilliottslab. A proteogenomic atlas of the human liver was generated using sc/snRNA-seq and CITE-seq on 19 liver biopsies, and 4 patients for Visium from ref. [15]. UMAP of Visium spot zonation and cell origin, as well as Visium (Visium, 10X Genomics) zonation data of the selected target were generated using the above-mentioned web resource.

### Inference of cell-type involvement from single-cell RNA-sequencing data

We examined cell type-specific transcriptional injury response in MASH in ∼ 138,000 single-cell transcriptomes accessed from the scRNA-seq **GSE207889 dataset. Therefore, we** performed a new analysis of the expression of selected targets using single-cell RNA-sequencing (scRNA-seq) data from the GEO repository above mentioned. We extracted pre-processed expression profiles for liver cell types (138053 cells x 31811 genes) modelled in HLOs developed from hPSCs as described by Hess et al. [16] and accessed from **GSE207889**.

The 3D culture model recapitulates human steatohepatitis through treatment with oleic acid (OA) and palmitic acid (PA) and fibrotic injury through treatment with transforming growth factor beta 1 (TGF-β1). Validation of cell identity performed as explained in ref. [16]. Cell type annotation was followed according to ScType database annotation method as used in the accessed data from **GSE207889.**

Challenge with OA induced steatosis and mild inflammatory changes in hepatocyte progenitors; challenge with PA induced a robust steatohepatitis signature from more mature hepatocyte subpopulations in the model [16] [16]. Single-cell RNA-sequencing (scRNA-seq) using Illumina NovaSeq 6000 (Homo sapiens) was used to investigate cell-type diversity. The experimental conditions included hPSC-derived HLOs cultured on an orbital shaker (OS) for 21 days and subsequently stimulated with TGF-β1 (10 ng/ml; R&D Systems 240-B-002), OA (OA-500 µM; Sigma Aldrich O1383) and PA (500 µM; Sigma Aldrich P0500) for four days and subjected to scRNAseq at day 25.

We reprocessed the information to ensure that the analysis included a sufficiently robust number of cells. Hence, the accessed file was subjected to quality control filtering and preprocessing using the Scampy Python package. This involved the removal of cells with less than 500 genes and genes expressed in less than 10 cells, which resulted in a total of 122,728 cells and 23,742 genes. Additionally, cells are filtered to ensure that the mitochondrial and ribosomal counts align with the 1.5 interquartile range of the respective data distribution. To prevent the inclusion of empty droplets in the cell count, cells with a doublet score greater than 0.5 (computed with Scrublet) were filtered out. This resulted in a gene expression matrix comprising 116,394 cells and 23,742 genes. Further steps include the normalisation and logarithmisation of the data. From the normalised matrix, we select the cells (82467 cells) including seven experimental conditions, comprising four controls and three treatments (denoted by “OA500”, “PA500” and “TGFB1”). In the subsequent stages, the three treatment conditions were analysed in relation to their respective control. The ForceAtlas2 algorithm [17] was used for the visualization of cell data in low-dimensional space.

To characterise the cells in the dataset, we computed the activity of protein-protein interaction networks (PPINs) derived from the scRNA-seq data. These networks are associated with several biological processes. In a nutshell, a PPIN associated with a biological process (or with a specific set of genes) can be constructed by considering the known interactions between proteins encoded by the genes within the defined gene set. A score that reflects the activity of a given PPIN can be defined for each cell by employing the methodology introduced by Senra et al. [18].

### Pathway analysis

A total of 572 biological processes (BPs) were computationally analysed using the QuickGO database [19]. This number was derived by considering BPs whose number of genes in the gene set ranged between 11 and 264 genes. To compute the PPIN activity for each BP in each cell, a reduced expression matrix of 5000 highly variable genes was considered. To analyse the TGFB1 and OA500 treatments, we have considered BPs correlated with the *ADGRG1* expression registered in all cells of *TGFB1*. To analyse the PA treatment, we consider BPs with a correlation of greater than 0.18 with *ADGRG1* expression, which is registered in all cells of this treatment. These correlations can be found in the **Supplementary Tables 4-6**. In addition to this functional analysis, we also perform a comparison of the expression levels of ADGRG1. This comparison is made between the treatment conditions and their respective controls on several populations of cell types. To achieve this, the Mann-Whitney test was employed.

### ADGRG1 tissue and hepatocyte interactions

The HumanBase, a collection of thousands of transcripts encoding diverse interaction data such as regulatory relationships, co-expression and protein-protein interactions is available at https://hb.flatironinstitute.org/gene/. This database, which applies machine learning algorithms and data-driven associations to learn biological associations from massive genomic data collections, was used to predict ADGRG1 liver tissue and hepatocyte interactions. Prediction uses genome-wide functional interaction networks for human tissues and cell types developed using a data-driven Bayesian methodology [20].

## Supplementary Tables

**Supplementary Table 1:**
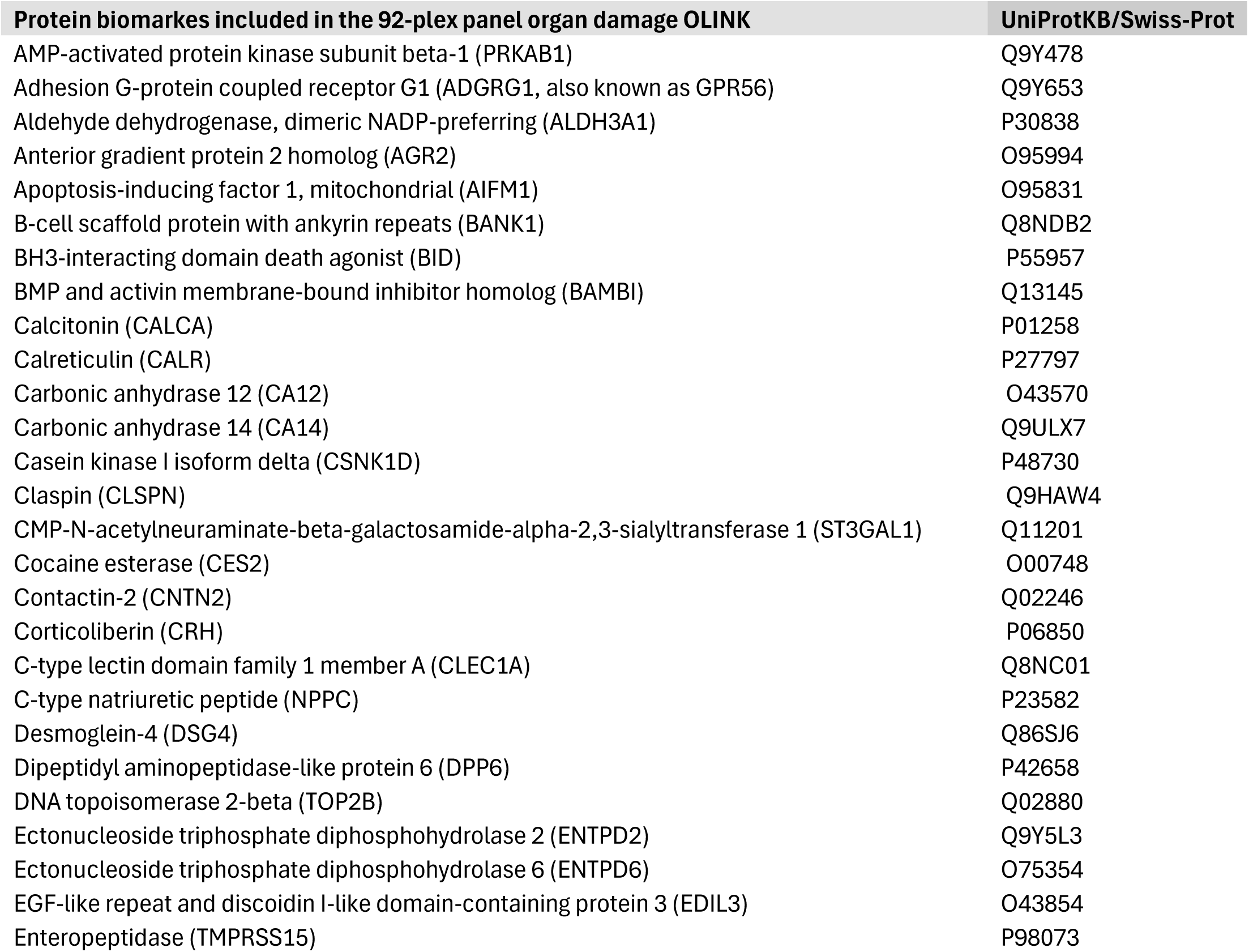
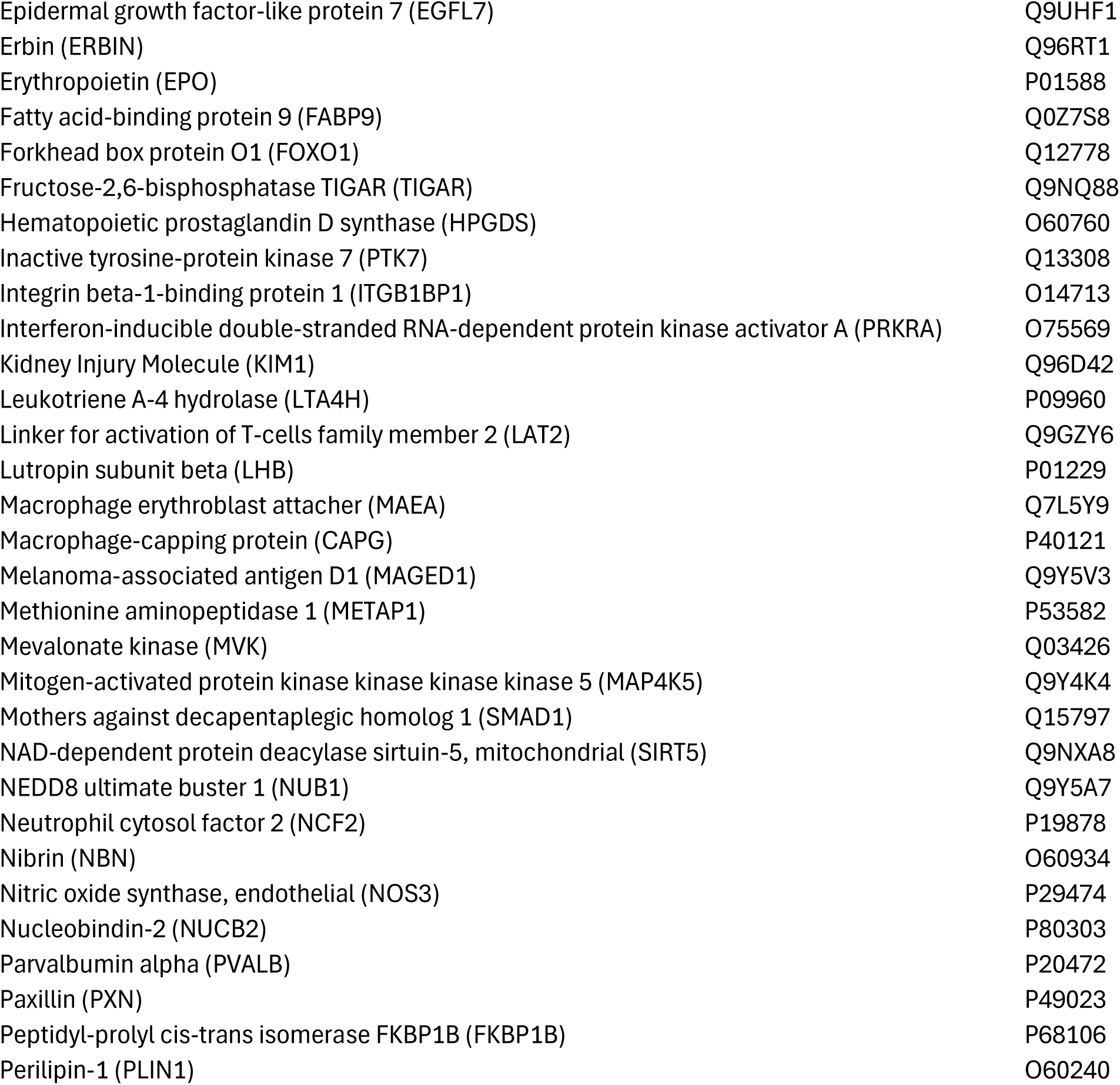
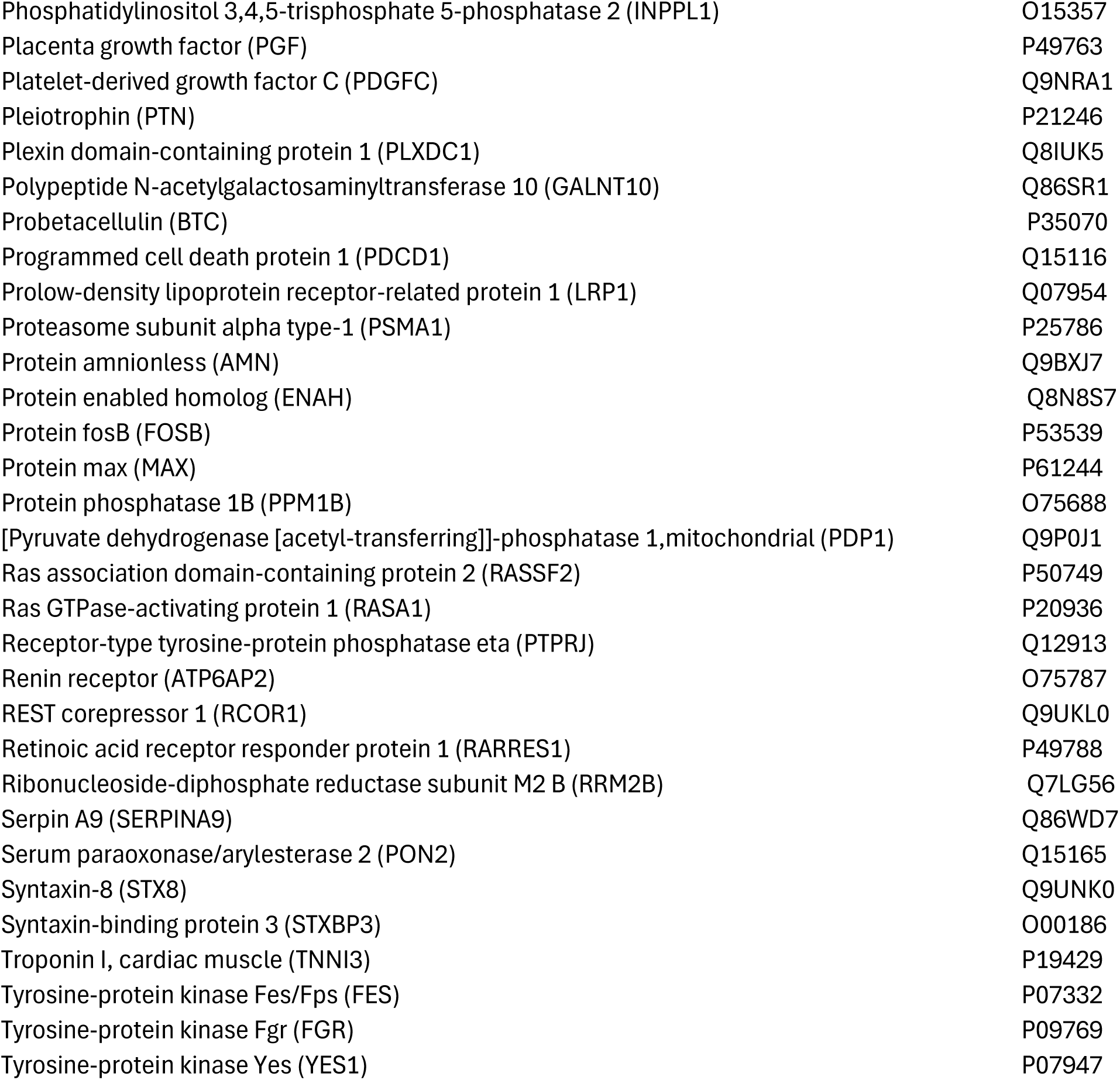
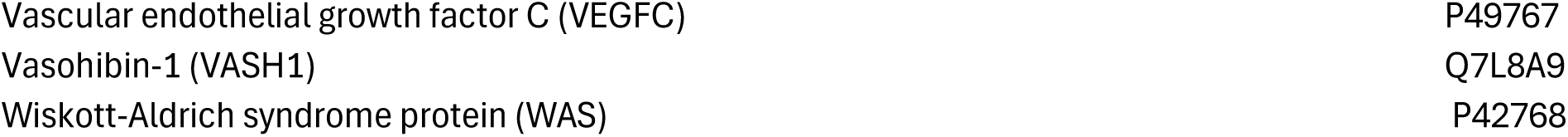
high throughput detection of prioritized signatures with pathophysiological importance in organ damage.

**Supplementary Table 2:**
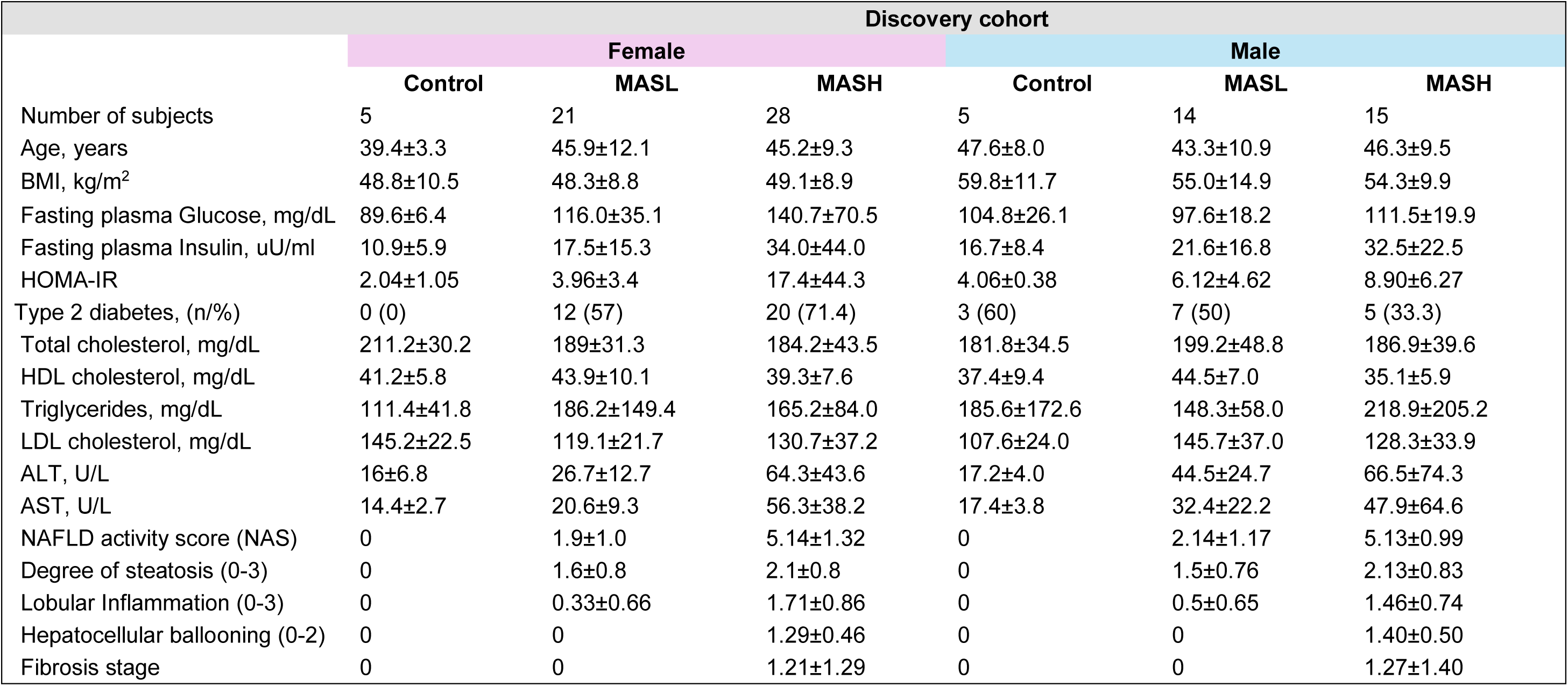
Self-reported gender-disaggregated demographic, clinical, biochemical, and histological characteristics of the patients included in the discovery phase.

|  | Discovery cohort |  |  |  |  |  |
| --- | --- | --- | --- | --- | --- | --- |
|  | Female |  |  | Male |  |  |
|  | Control | MASL | MASH | Control | MASL | MASH |
| Number of subjects | 5 | 21 | 28 | 5 | 14 | 15 |
| Age, years | 39.4±3.3 | 45.9±12.1 | 45.2±9.3 | 47.6±8.0 | 43.3±10.9 | 46.3±9.5 |
| BMI, kg/m<sup>2</sup> | 48.8±10.5 | 48.3±8.8 | 49.1±8.9 | 59.8±11.7 | 55.0±14.9 | 54.3±9.9 |
| Fasting plasma Glucose, mg/dL | 89.6±6.4 | 116.0±35.1 | 140.7±70.5 | 104.8±26.1 | 97.6±18.2 | 111.5±19.9 |
| Fasting plasma Insulin, uU/ml | 10.9±5.9 | 17.5±15.3 | 34.0±44.0 | 16.7±8.4 | 21.6±16.8 | 32.5±22.5 |
| HOMA-IR | 2.04±1.05 | 3.96±3.4 | 17.4±44.3 | 4.06±0.38 | 6.12±4.62 | 8.90±6.27 |
| Type 2 diabetes, (n/%) | 0 (0) | 12 (57) | 20 (71.4) | 3 (60) | 7 (50) | 5 (33.3) |
| Total cholesterol, mg/dL | 211.2±30.2 | 189±31.3 | 184.2±43.5 | 181.8±34.5 | 199.2±48.8 | 186.9±39.6 |
| HDL cholesterol, mg/dL | 41.2±5.8 | 43.9±10.1 | 39.3±7.6 | 37.4±9.4 | 44.5±7.0 | 35.1±5.9 |
| Triglycerides, mg/dL | 111.4±41.8 | 186.2±149.4 | 165.2±84.0 | 185.6±172.6 | 148.3±58.0 | 218.9±205.2 |
| LDL cholesterol, mg/dL | 145.2±22.5 | 119.1±21.7 | 130.7±37.2 | 107.6±24.0 | 145.7±37.0 | 128.3±33.9 |
| ALT, U/L | 16±6.8 | 26.7±12.7 | 64.3±43.6 | 17.2±4.0 | 44.5±24.7 | 66.5±74.3 |
| AST, U/L | 14.4±2.7 | 20.6±9.3 | 56.3±38.2 | 17.4±3.8 | 32.4±22.2 | 47.9±64.6 |
| NAFLD activity score (NAS) | 0 | 1.9±1.0 | 5.14±1.32 | 0 | 2.14±1.17 | 5.13±0.99 |
| Degree of steatosis (0-3) | 0 | 1.6±0.8 | 2.1±0.8 | 0 | 1.5±0.76 | 2.13±0.83 |
| Lobular Inflammation (0-3) | 0 | 0.33±0.66 | 1.71±0.86 | 0 | 0.5±0.65 | 1.46±0.74 |
| Hepatocellular ballooning (0-2) | 0 | 0 | 1.29±0.46 | 0 | 0 | 1.40±0.50 |
| Fibrosis stage | 0 | 0 | 1.21±1.29 | 0 | 0 | 1.27±1.40 |

**Supplementary Table 3:**
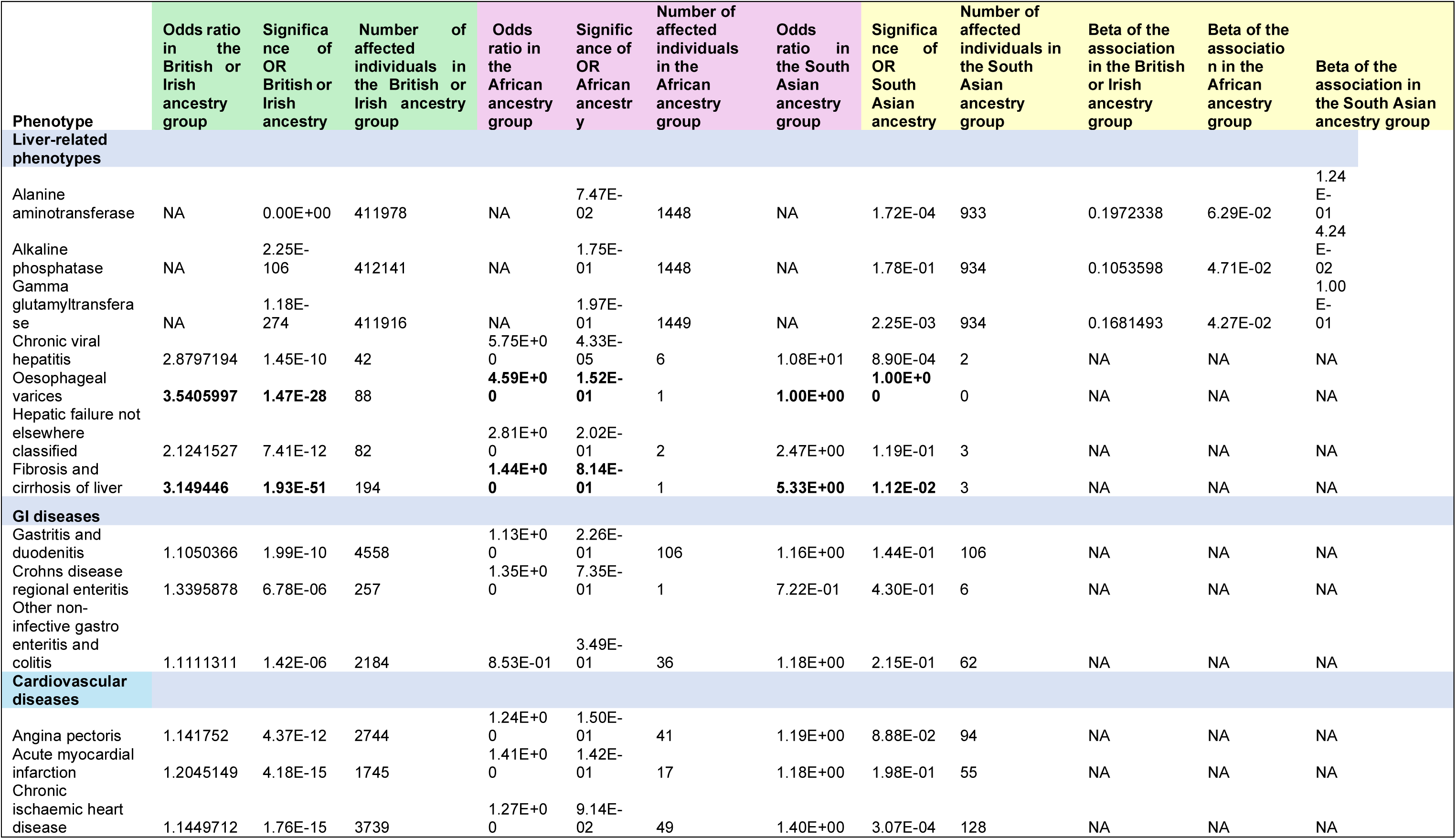
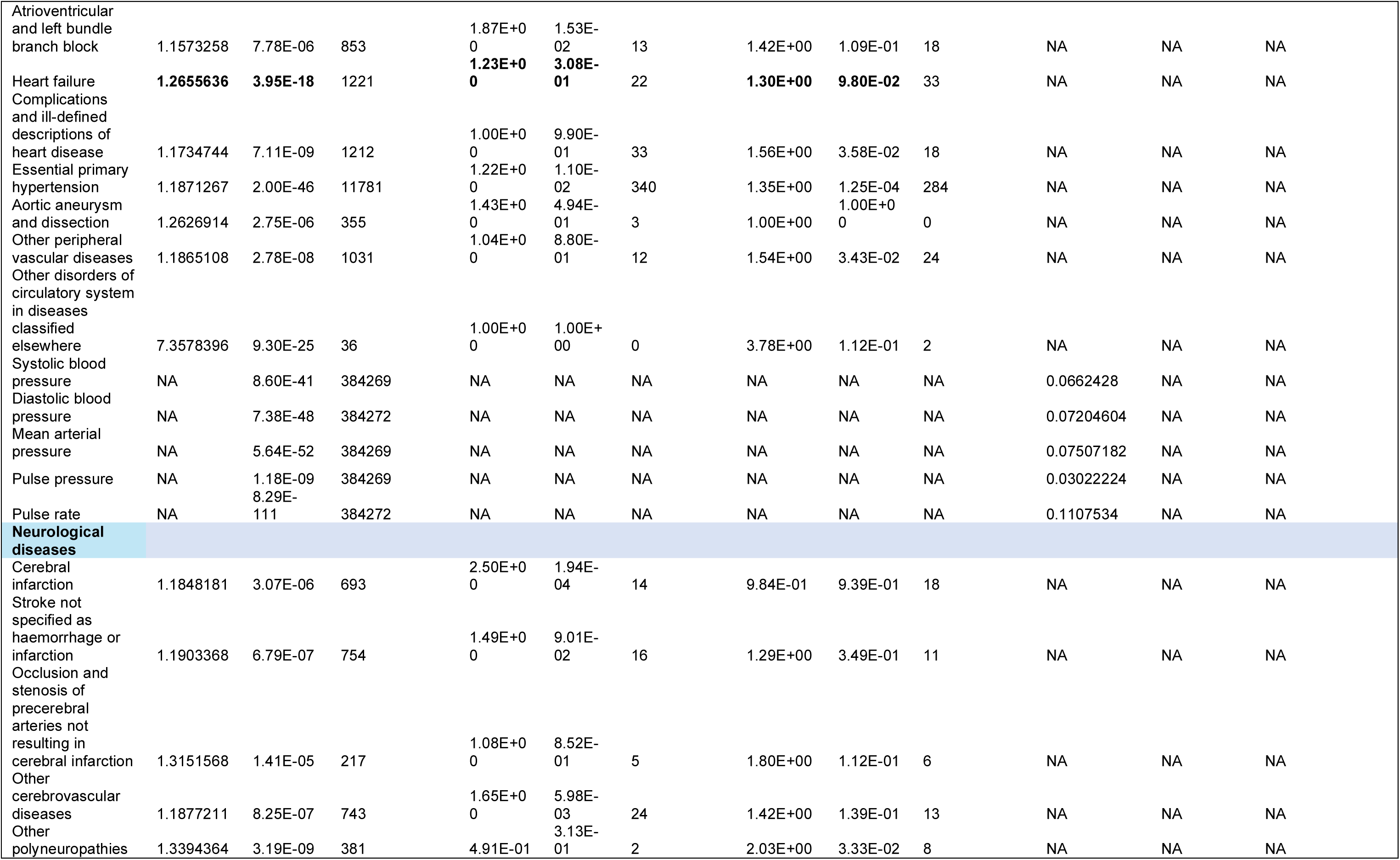
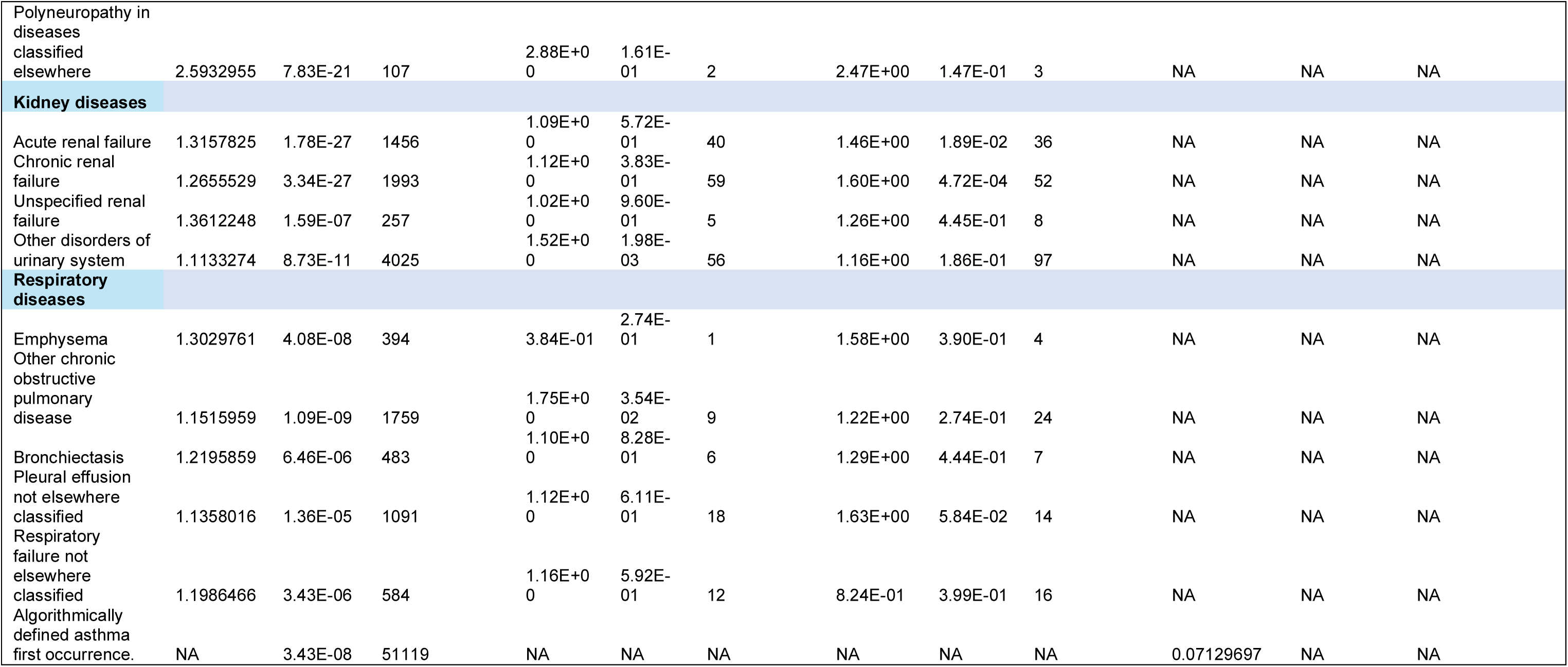
ADGRG1 protein-phenotype associations in the UKBB population stratified on ancestry.

**Supplementary Table 4:** Enriched gene ontology (GO) biological pathways associated with liver ADGRG1 expression in response to the inducing stimulus OA-500 mM.

| Biological process OA-500 mM day 25 OS-HLOs |  |  |  |
| --- | --- | --- | --- |
| BP GOID | BP GO Term | P-value | Correlation with ADGRG1 |
| GO:0001570 | vasculogenesis | 3.77236E-10-23 | 0.23557 |
| GO:0071604 | transforming growth factor beta production | 1.21488E-10-6 | 0.223742 |
| GO:0097485 | neuron projection guidance | 3.48338E-10-7 | 0.215978 |
| GO:0030216 | keratinocyte differentiation | 4.05969E-05 | 0.208908 |
| GO:0009913 | epidermal cell differentiation | 3.00681E-05 | 0.208583 |
| GO:0006402 | mRNA catabolic process | 5.76938E-10-6 | 0.204078 |
| GO:0042100 | B cell proliferation | 4.3279E-10-6 | 0.203815 |
| GO:0060070 | canonical Wnt signaling pathway | 1.72009E-10-8 | 0.202825 |
| GO:0060389 | pathway-restricted SMAD protein phosphorylation | 2.82739E-10-11 | 0.202271 |
| GO:0048041 | focal adhesion assembly | 1.14078E-10-8 | 0.202014 |
| GO:0031424 | keratinization | 3.34608E-05 | 0.201676 |
| GO:0016311 | dephosphorylation | 8.59084E-05 | 0.201624 |
| GO:0044774 | mitotic DNA integrity checkpoint signaling | 9.07961E-05 | 0.198733 |
| GO:0031570 | DNA integrity checkpoint signaling | 7.95562E-05 | 0.198281 |
| GO:0030225 | macrophage differentiation | 1.45779E-05 | 0.198112 |
| GO:0042770 | signal transduction in response to DNA damage | 1.83426E-05 | 0.197758 |
| GO:0033627 | cell adhesion mediated by integrin | 4.57397E-10-21 | 0.196732 |
| GO:0010761 | fibroblast migration | 6.06922E-10-6 | 0.195652 |
| GO:0060317 | cardiac epithelial to mesenchymal transition | 5.15895E-10-10 | 0.195596 |
| GO:0035987 | endodermal cell differentiation | 1.32987E-10-17 | 0.195478 |
| GO:0043534 | blood vessel endothelial cell migration | 3.26947E-10-9 | 0.194335 |
| GO:0071542 | dopaminergic neuron differentiation | 5.61804E-05 | 0.189806 |
| GO:0071897 | DNA biosynthetic process | 0.017547 | 0.189473 |
| GO:0007405 | neuroblast proliferation | 0.000186782 | 0.188924 |
| GO:0006470 | protein dephosphorylation | 0.00032841 | 0.188917 |
| GO:0002062 | chondrocyte differentiation | 7.75761E-10-8 | 0.187484 |
| GO:0045109 | intermediate filament organization | 3.46285E-10-6 | 0.187218 |
| GO:0001837 | epithelial to mesenchymal transition | 1.49512E-10-8 | 0.186729 |
| GO:1904019 | epithelial cell apoptotic process | 1.36062E-10-13 | 0.185506 |
| GO:0046700 | heterocycle catabolic process | 0.000114627 | 0.185393 |
| GO:0034976 | response to endoplasmic reticulum stress | 9.36847E-10-6 | 0.185107 |
| GO:0016197 | endosomal transport | 1.29644E-05 | 0.184475 |
| GO:0002244 | hematopoietic progenitor cell differentiation | 1.53827E-10-7 | 0.183458 |
| GO:0048008 | platelet-derived growth factor receptor signaling pathway | 0.000684402 | 0.180356 |

**Supplementary Table 5:** Enriched gene ontology (GO) biological pathways associated with liver ADGRG1 expression in response to the inducing stimulus PA-500 mM.

| Biological process PA-500 mM day 25 OS-HLOs |  |  |  |
| --- | --- | --- | --- |
| BP GOID | BP GO Term | P-value | Correlation with ADGRG1 |
| GO:0071604 | transforming growth factor beta production | 6.92563E-10-7 | 0.185996 |
| GO:0010573 | vascular endothelial growth factor production | 1.95964E-10-33 | 0.188373 |
| GO:0090077 | foam cell differentiation | 6.45197E-10-39 | 0.180614 |
| GO:0034113 | heterotypic cell-cell adhesion | 7.59492E-10-20 | 0.186238 |
| GO:0032637 | interleukin-8 production | 2.88137E-10-14 | 0.183184 |
| GO:0033627 | cell adhesion mediated by integrin | 1.79989E-10-49 | 0.199759 |
| GO:0001570 | vasculogenesis | 7.10424E-10-27 | 0.187717 |
| GO:0014902 | myotube differentiation | 1.28469E-10-15 | 0.193003 |
| GO:0007229 | integrin-mediated signaling pathway | 1.40715E-10-55 | 0.182477 |
| GO:0097193 | intrinsic apoptotic signaling pathway | 8.73888E-10-9 | 0.182816 |
| GO:0042180 | cellular ketone metabolic process | 5.01193E-05 | 0.188218 |

**Supplementary Table 6:** Enriched gene ontology (GO) biological pathways associated with liver ADGRG1 expression in response to the inducing stimulus TGFB1-10ng.

| **Biological process TGFB1-10ng day 25 OS-HLOs** |  |  |  |
| --- | --- | --- | --- |
| **BP GOID** | **BP GO Term** | **P-value** | **Correlation with ADGRG1** |
| GO:0030216 | keratinocyte differentiation | 5.46205E-10-109 | 0.247736 |
| GO:0009913 | epidermal cell differentiation | 4.29707E-10-110 | 0.247614 |
| GO:0031424 | keratinization | 4.11108E-10-90 | 0.243944 |
| GO:0150115 | cell-substrate junction organization | 2.34867E-10-90 | 0.217439 |
| GO:0030855 | epithelial cell differentiation | 1.24585E-10-86 | 0.209131 |
| GO:0099536 | synaptic signaling | 1.19711E-10-108 | 0.20527 |
| GO:0034446 | substrate adhesion-dependent cell spreading | 1.55737E-10-80 | 0.205258 |
| GO:0045109 | intermediate filament organization | 1.61365E-10-83 | 0.204331 |
| GO:0045103 | intermediate filament-based process | 2.52106E-10-97 | 0.201534 |
| GO:0086003 | cardiac muscle cell contraction | 6.49171E-10-64 | 0.196075 |
| GO:0086065 | cell communication involved in cardiac conduction | 2.31505E-10-63 | 0.195554 |
| GO:0086002 | cardiac muscle cell action potential involved in contraction | 5.81261E-10-63 | 0.195153 |
| GO:0034113 | heterotypic cell-cell adhesion | 1.20102E-10-58 | 0.191508 |
| GO:0034109 | homotypic cell-cell adhesion | 1.61067E-10-58 | 0.18281 |
| GO:0030099 | myeloid cell differentiation | 1.19766E-10-63 | 0.182499 |
| GO:0048864 | stem cell development | 1.21651E-10-60 | 0.180761 |
| GO:0014033 | neural crest cell differentiation | 2.60967E-10-61 | 0.18032 |
| GO:0051604 | protein maturation | 2.69864E-10-85 | 0.175244 |
| GO:0048863 | stem cell differentiation | 1.40985E-10-55 | 0.17046 |
| GO:0043113 | receptor clustering | 1.4505E-10-53 | 0.16895 |
| GO:0002065 | columnar/cuboidal epithelial cell differentiation | 6.55974E-10-46 | 0.168365 |
| GO:0034620 | cellular response to unfolded protein | 9.28871E-10-30 | 0.168332 |
| GO:0035967 | cellular response to topologically incorrect protein | 9.28871E-10-30 | 0.168332 |
| GO:1904019 | epithelial cell apoptotic process | 1.42686E-10-43 | 0.168328 |
| GO:0051259 | protein complex oligomerization | 6.76531E-10-49 | 0.164617 |
| GO:0051262 | protein tetramerization | 6.09318E-10-49 | 0.164607 |
| GO:0070371 | ERK1 and ERK2 cascade | 1.5278E-10-56 | 0.163555 |
| GO:0072659 | protein localization to plasma membrane | 3.11803E-10-60 | 0.160988 |
| GO:0002064 | epithelial cell development | 1.52609E-10-35 | 0.159117 |
| GO:0033627 | cell adhesion mediated by integrin | 8.90619E-10-32 | 0.158654 |
| GO:0014065 | phosphatidylinositol 3-kinase signaling | 3.9896E-10-45 | 0.158253 |
| GO:0001909 | leukocyte mediated cytotoxicity | 1.62084E-10-36 | 0.157811 |
| GO:0006886 | intracellular protein transport | 4.54784E-10-44 | 0.156639 |
| GO:0019221 | cytokine-mediated signaling pathway | 1.24439E-10-15 | 0.156479 |
| GO:0008625 | extrinsic apoptotic signaling pathway via death domain receptors | 2.78617E-10-32 | 0.156127 |
| GO:0009267 | cellular response to starvation | 1.16648E-10-31 | 0.15219 |
| GO:0002573 | myeloid leukocyte differentiation | 5.58344E-10-34 | 0.151508 |
| GO:0051169 | nuclear transport | 6.99881E-10-33 | 0.150173 |

